# A conserved cargo recognition mechanism establishes TANGO1 as a selective extracellu lar matrix cargo receptor

**DOI:** 10.64898/2026.07.20.26358213

**Authors:** Julian Nüchel, Andrea Accogli, Steffen Lütke, Sonja Titze, Sabine Wilhelm, Marina Tauber, Franziska Oehlert, Manuel Koch, Valeria Capra, Reza Maroofian, Mina Zamani, Mateja Vinkšel Prah, Karen E. Heath, Agnes Bloch-Zupan, Amaka C. Offiah, TANGO1 consortium, Markus Plomann, Beate Eckes, Thomas Krieg, Daniel Kümmel, Alvise Schiavinato, Gerhard Sengle, Constantinos Demetriades

**Affiliations:** University of Cologne, Faculty of Medicine and University Hospital Cologne, Center for Biochemistry, 50931 Cologne, Germany; Max Planck Institute for Biology of Ageing (MPI-AGE), 50931 Cologne, Germany; Center for Molecular Medicine Cologne (CMMC), University of Cologne, 50931 Cologne, Germany; Division of Medical Genetics, Department of Medicine, McGill University Health Center, Montreal, QC, Canada; Child Health and Human Development Program - Research Institute of the McGill University Health Centre, Montreal, Québec, Canada; Department of Human Genetics, McGill University, Montreal, QC, Canada; University Hospital Cologne, Department of Pediatrics and Adolescent Medicine, 50931 Cologne, Germany; University of Münster, Institute of Biochemistry, 48149 Münster, Germany; University of Cologne, Institute for Dental Research and Oral Musculoskeletal Biology, Faculty of Medicine and University Hospital Cologne, 50931 Cologne, Germany; Center for Dental, Oral and Maxillofacial Medicine (Central Facilities), Faculty of Medicine and University Hospital Cologne, 50931 Cologne, Germany; Genomic and Clinical Genetics Unit, IRCSS Istituto G. Gaslini, Genoa, Italy; Department of Neuromuscular Diseases, Queen Square Institute of Neurology, University College London, London, UK; Department of Biology, New York University, New York, NY, USA; University Medical Centre Ljubljana, Clinical Institute of Genomic Medicine, Ljubljana, Slovenia; Institute of Medical and Molecular Genetics (INGEMM), Hospital Universitario La Paz, IdiPAZ, UAM, Madrid, Spain; Skeletal dysplasia multidisciplinary Unit (UMDE-ERN BOND), Hospital Universitario la Paz, Madrid, Spain; CIBERER, ISCIII, Madrid, Spain; Faculté de Chirurgie Dentaire Robert Frank, Université de Strasbourg, Strasbourg 67000, France; Centre de Référence des maladies rares orales et dentaires, Reference Center for Rare Oral and Dental Diseases, CRMR-O-Rares, Hôpitaux Universitaires de Strasbourg (HUS), Filière de Santé Maladies Rares TETECOU, European Reference Network ERN CRANIO, France; Institute of Genetics and Molecular and Cellular Biology, INSERM U1258, CNRS UMR7104, Université de Strasbourg, Illkirch-Graffenstaden 67400, France; Chair of Paediatric Musculoskeletal Imaging and Honorary Consultant Paediatric Radiologist, University of Sheffield, Sheffield, UK; University of Cologne, Faculty of Medicine and University Hospital Cologne, Translational Matrix Biology, 50931 Cologne, Germany; University of Cologne, Cologne Excellence Cluster on Cellular Stress Responses in Aging-Associated Diseases (CECAD), 50931 Cologne, Germany; Cologne Center for Musculoskeletal Biomechanics (CCMB), 50931 Cologne, Germany; European Research Institute for the Biology of Ageing (ERIBA), University of Groningen (RUG), University Medical Center Groningen (UMCG), 9713 GZ Groningen, The Netherlands

**Author notes:** Correspondence (J.N.); (A.A.); (C.D.); (C.D.). Individual members and their affiliations appear in Supplementary Information (TANGO1 consortium). Dr. Plomann has sadly passed away during preparation of the manuscript.

**Keywords:** TANGO1, *MIA3*, ECM secretion, cargo recognition, ER exit sites, collagen trafficking, skeletal dysplasia

## Abstract

The endoplasmic reticulum (ER) orchestrates the secretion of extracellular matrix (ECM) proteins, many of which exceed the size of conventional transport vesicles and therefore require specialized export machinery. TANGO1, encoded by *MIA3*, organizes ER exit sites for bulky cargo export, yet the molecular basis of cargo recognition and its relationship to the collagen-specific chaperone HSP47 remain unclear. Through quantitative secretome profiling of TANGO1- and HSP47-deficient fibroblasts, structural modeling, and binding analyses, we show that TANGO1 directly and selectively recognizes a defined subset of ECM proteins, including specific collagen and fibrillin isoforms, through a conserved tyrosine residue in its luminal MOTH domain, independently of HSP47. Novel *MIA3* variants identified in individuals with previously undescribed skeletal dysplasia disrupt this cargo-binding interface, leading to selective intracellular retention of ECM proteins. These findings identify direct cargo recognition, rather than ER exit-site assembly, as the primary molecular defect underlying *MIA3*/TANGO1-associated skeletal dysplasia. More broadly, our work establishes TANGO1 as a selective ECM cargo receptor that functions independently of HSP47, providing a new framework for understanding bulky cargo selection at the ER.

## Introduction

The endoplasmic reticulum (ER) coordinates synthesis, folding, and export of secretory proteins. While conventional coat protein complex II (COPII) vesicles measure 60-80 nm in diameter, many extracellular matrix (ECM) proteins, including collagens, fibrillins, and proteoglycans, exceed this capacity due to their size, rigidity and complex supramolecular organization. The export of such cargo therefore requires specialized gatekeepers that control selective passage through ER exit sites (ERES) ^1,2^. Collagen assembly in the ER involves the formation of defined heterotrimeric complexes, such as collagen VI, in which individual chains fulfill non-redundant roles ^3^. This raises the possibility that selective handling of specific ECM chains may be required during early secretory pathway steps.

TANGO1 (Transport and Golgi Organization 1) has emerged as a central factor in bulky cargo export. Localized at ERES, TANGO1 interacts with COPII components and recruits membranes from the ER-Golgi intermediate compartment (ERGIC), facilitating formation of enlarged carriers ^4,5^. Although initially described as a collagen export factor, subsequent studies implicated TANGO1 in the secretion of additional ECM proteins ^4–7^. Importantly, however, it remains unclear whether TANGO1 acts as a generic ECM secretion factor or via selective cargo recognition.

Despite extensive investigation, several fundamental questions regarding TANGO1 function remain unresolved. First, it is unclear whether TANGO1 directly recognizes and binds its cargo within the ER lumen, or whether it primarily serves as a structural scaffold at ERES that indirectly facilitates export. Second, although TANGO1 has been implicated in the secretion of large ECM proteins, it remains uncertain whether cargo size alone determines TANGO1 dependency. Third, the functional relationship between TANGO1 and the collagen-specific chaperone HSP47 is incompletely defined, and it is not known whether TANGO1 requires HSP47 to mediate cargo recognition and export. HSP47 has previously been described to be required for stabilization of the collagen triple helical structure ^8,9^, thereby preventing misfolding and aggregation. Furthermore, while HSP47 mediates cargo recognition and secretion of collagen type I, this is not the case for collagen type VI ^10^. Previous interaction studies proposed that TANGO1 does not directly bind collagen but instead engages cargo through HSP47 that functions as an adaptor ^8,11^. More recent structural analyses of the luminal SH3-like/MOTH (MIA, Otoraplin, TALI homology) domain (for simplicity, hereafter referred to as MOTH) domain suggest potential direct binding capabilities ^12^, yet cargo selectivity remains incompletely defined. Moreover, whether TANGO1 functions as a universal mediator of large cargo export or instead discriminates among structurally related ECM proteins is unknown.

In addition to its role in ECM secretion, genetic studies have highlighted the physiological importance of TANGO1 in human development. Pathogenic variants in *MIA3*, the gene encoding TANGO1, have been associated with severe skeletal disorders characterized by impaired connective tissue formation. For example, biallelic variants affecting the cytosolic region of TANGO1 have been associated with skeletal dysplasia and dentinogenesis imperfecta, indicating that disruption of TANGO1-mediated secretion can profoundly affect bone and tooth development ^13,14^. Consistently, complete loss of TANGO1 function in animal models leads to embryonic lethality, further underscoring its essential role in organismal development ^13^. More recently, a missense variant (p.Cys38Phe; hereafter C38F) within the luminal MOTH domain of TANGO1 has been reported in association with severe developmental phenotypes, although the mechanistic consequences of this mutation remain poorly understood ^15^.

Here, we show that TANGO1 acts as a selective cargo receptor that directly binds specific ECM chains via its MOTH domain, independently of HSP47. Through secretome analysis of patient-derived TANGO1/*MIA3* MOTH domain variants and biochemical binding analyses, we uncover a cargo-selective mechanism of ECM export and define the molecular mechanisms of bulky cargo selection at ERES.

## Results

### Quantitative secretome profiling reveals selective, HSP47-independent cargo dependency on TANGO1

To systematically define the cargo spectrum of TANGO1 and to assess its functional relationship with the collagen chaperone HSP47, we performed quantitative SILAC-based secretome analysis ^16^ in WI-26 human diploid lung fibroblasts lacking either TANGO1 or HSP47 (Figure 1a). Two independent CRISPR/Cas9-derived knockout clones for each gene were compared with matched control cells in order to ensure reproducibility and exclude clone-specific effects. Analysis of conditioned media from TANGO1-deficient cells revealed a pronounced reduction in the secretion of 171 proteins relative to wild-type controls (Figure 1b,c and Supplementary Table 1). Gene ontology enrichment analysis of the downregulated secreted proteins showed a strong overrepresentation of factors associated with ECM organization and collagen maturation, consistent with the proposed role of TANGO1 in the export of bulky ECM cargo. Among the most strongly affected proteins were large structural ECM components including COL1A2, COL12A1, COL6A3, and fibrillin-2 (FBN2), supporting that TANGO1 is required for efficient secretion of multiple matrix proteins.

**Figure 1.**
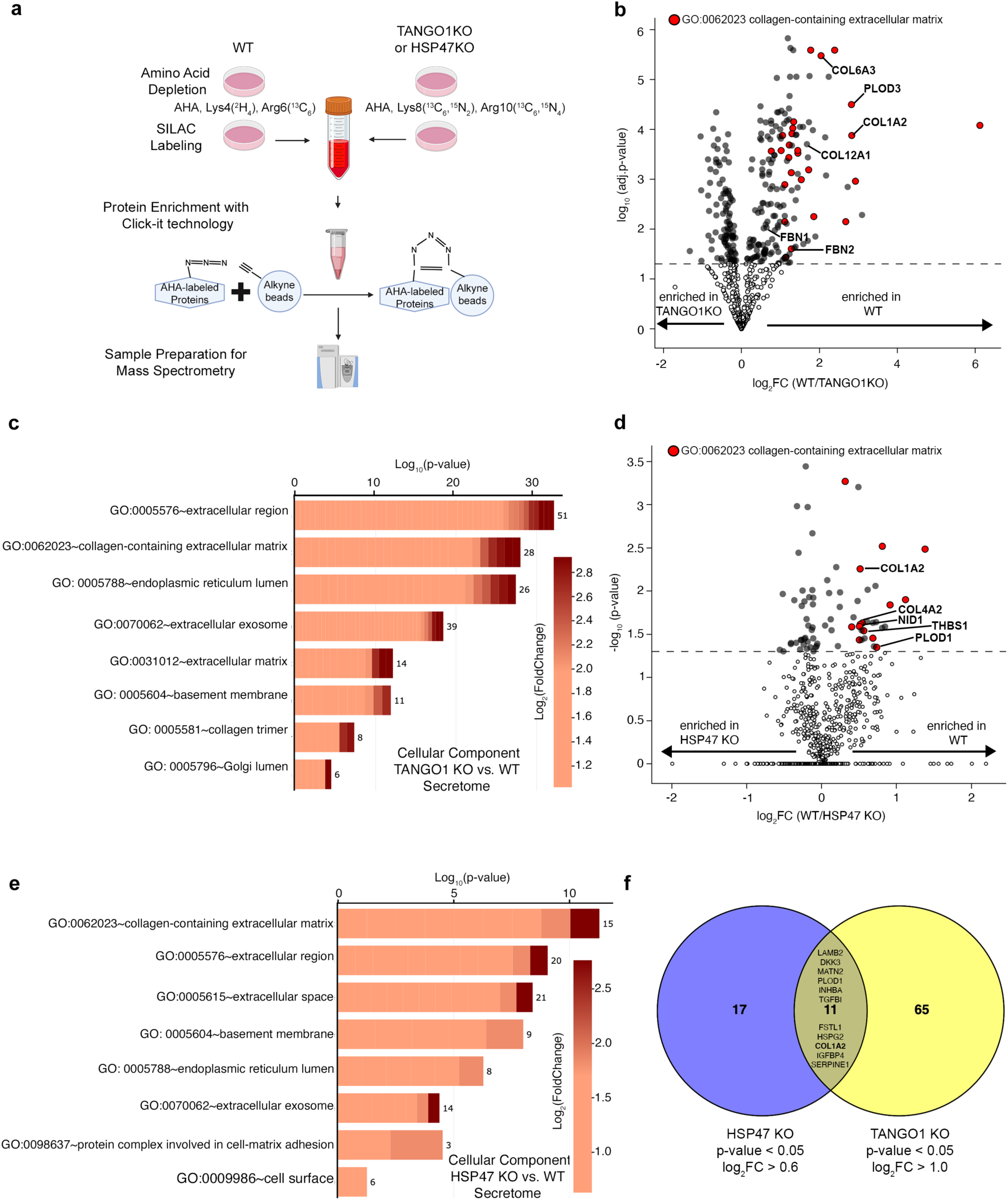
Secretion of selected ECM cargo is affected in TANGO1-deficient cells, independent of HSP47. **(a)** Experimental outline of the SILAC labeling- and click chemistry-based assay performed to identify the TANGO1 or HSP47-dependent secretome in WI-26 cells (see also Methods). **(b-c)** Volcano plot in (b) showing all proteins identified in the TANGO1-dependent secretome (gray dots). Proteins whose secretion is strongly and significantly decreased (Log_2_FC>1.0, p<0.05) in TANGO1 KO WI-26 cells were used for the GO analysis shown in (c). Proteins within this subset that belong to the cellular component (BP) GO term ‘collagen-containing extracellular matrix’ are shown in red. **(d-e)** Volcano plot in (d) showing all proteins identified in the HSP47-dependent secretome (gray dots). Proteins whose secretion is strongly and significantly decreased (Log_2_FC>0.6, p<0.05) in HSP47 KO WI-26 cells were used for the GO analysis shown in (e). Proteins within this subset that belong to the cellular component (BP) GO term ‘collagen-containing extracellular matrix’ are shown in red. (f) Venn diagram comparing the TANGO1 and HSP47-dependent secreted proteome from the two SILAC datasets.

Strikingly, closer inspection of this dataset revealed a striking degree of cargo selectivity. Within the same collagen families, individual chains exhibited markedly different dependencies on TANGO1. For example, secretion of COL6A3 was strongly reduced in TANGO1 knockout cells, whereas secretion of the structurally related COL6A1 chain was only modestly affected. A similar pattern was observed for type I collagen chains, where COL1A2 secretion was strongly diminished while COL1A1 secretion remained largely preserved. Comparable selectivity was also observed among non-collagenous ECM proteins: secretion of FBN2 was substantially impaired in the absence of TANGO1, whereas secretion of the closely related fibrillin-1 (FBN1) was largely unaffected. These findings indicate that TANGO1 does not function as a universal mediator of bulky cargo export but instead discriminates among structurally related ECM chains. In addition to structural matrix proteins, the TANGO1-dependent secretome included several enzymes involved in collagen modification and maturation, such as LOXL2 and the prolyl- and lysyl-hydroxylases P4HA1 and PLOD1-3, which are known to be essential for the formation of stable collagen crosslinks ^17–19^. Reduced secretion of these factors suggests that loss of TANGO1 may affect not only the abundance but also the intrinsic stability of the ECM.

Since it was previously proposed that TANGO1 recognizes collagen cargo through interaction with the ER-resident chaperone HSP47 ^8^, we next examined the secretome of HSP47-deficient cells under identical experimental conditions (Figure 1d,e and Supplementary Table 2). In contrast to TANGO1 knockout cells, loss of HSP47 resulted in a substantially smaller set of secretion defects, with 36 proteins significantly reduced in the extracellular medium. Gene ontology analysis again revealed enrichment of ECM components, but the affected proteins differed markedly from those observed in TANGO1 knockout cells. In particular, many of the HSP47-dependent proteins were associated with basement membrane organization, including COL4A2, NID1, and LAMB2 ^20^. Direct comparison of the two secretome datasets revealed only limited overlap between TANGO1-dependent and HSP47-dependent cargo (Figure 1f). Sixty-five proteins were uniquely dependent on TANGO1 but unaffected by loss of HSP47, including several large ECM proteins such as COL6A3, COL12A1, and FBN2. Conversely, only seventeen proteins were uniquely dependent on HSP47. Notably, COL1A2 represented the only collagen chain that showed significant dependency on both proteins. These findings indicate that TANGO1-mediated export of several ECM cargo proteins occurs independently of HSP47 and suggest that the two proteins fulfill distinct roles within the early secretory pathway.

To validate the secretome analysis and to examine the intracellular fate of TANGO1-dependent cargo proteins, we performed immunoblot analyses of cell lysates and conditioned media (Figure 2a) derived from the same knockout cell lines used in the proteomic experiments. Consistent with the mass spectrometry results, secretion of several ECM proteins, including COL1A2 (Figure 2a,d,e), COL6A3 (Figure 2a,h,i), COL7 (Figure 2a,j,k), COL12 (Figure 2a,l,m), and FBN2 (Figure 2a,p,q), was markedly reduced in TANGO1-deficient cells. In contrast, secretion of COL1A1 (Figure 2a-c), COL6A1 (Figure 2a,f,g), and FBN1 (Figure 2a,n,o) remained largely unaffected. Strikingly, all ECM proteins analyzed showed increased intracellular accumulation in TANGO1 knockout cells, indicating that these proteins are synthesized but fail to exit the ER (Figure 2). This intracellular retention was observed even for cargo proteins whose secretion was only partially reduced, suggesting that loss of TANGO1 impairs ER export rather than protein synthesis or stability. In contrast, HSP47-deficient cells exhibited a different phenotype. Rather than accumulating intracellular collagen, these cells displayed reduced intracellular levels of several collagen chains, consistent with the established role of HSP47 as a folding chaperone that stabilizes the collagen triple helix (Figure 2a-o). Thus, while TANGO1 deficiency primarily results in ER retention of ECM cargo, HSP47 deficiency leads to reduced intracellular collagen abundance, likely reflecting impaired folding and increased degradation. Together, these observations demonstrate that TANGO1 and HSP47 regulate distinct steps in collagen biogenesis: TANGO1 mediates selective ER export, whereas HSP47 primarily supports collagen folding and stability.

**Figure 2.**
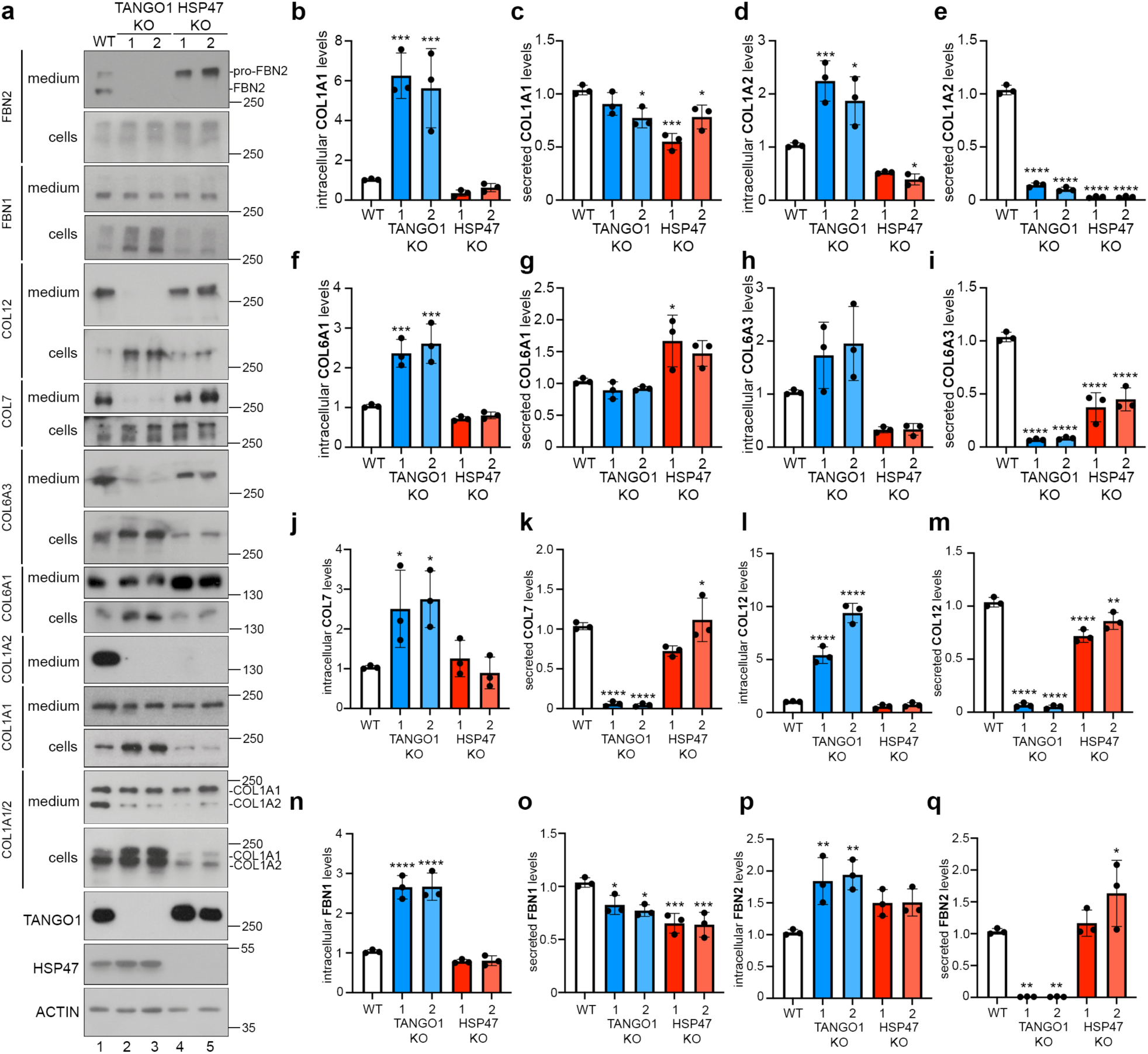
TANGO1, but not HSP47 is required for selective ER export of bulky ECM cargo proteins. **(a)** Western blot analysis of cell lysates (cells) and supernatants (media) of WT, two independent clones of TANGO1 KO cells and HSP47 KO cells, cultured for 24 h in the presence of ascorbic acid using the indicated antibodies. **(b-q)** Densiometric quantification of intracellular levels (b) and secreted levels (c) of type I collagen alpha 1 chain (COL1A1), intracellular (d) and secreted levels (e) of type I collagen alpha 2 chain (COL1A2). The intracellular levels of type VI collagen alpha 1 chain (COL6A1, f), of type VI collagen alpha 3 chain (COL6A3, h), of type VII collagen (COL7, j), of type XII collagen (COL12, l), fibrillin-1 (FBN1, n) and fibrillin-2 (FBN2, p) as well as the secreted levels of COL6A1 (g), COL6A3 (i), COL7 (k), COL12 (m), FBN1 (o) and FBN2 (q) are quantified. * p< 0.05, ** p< 0.01, *** p< 0.001, **** p<0.0001, one-way ANOVA. Data in graphs shown as mean ± SD. n = 3 independent experiments.

### Cells lacking TANGO1 or HSP47 exhibit specific ECM assembly defects

To determine whether the observed secretion defects translate into impaired ECM formation, we examined ECM assembly in confluent fibroblast cultures in the presence of ascorbic acid-2-phosphate. Immunofluorescence analysis revealed the formation of extensive fibrillar networks of collagen and fibrillin proteins in wild-type cells (Figure 3a-f). In contrast, TANGO1 knockout cells failed to assemble normal ECM networks. Fibers composed of COL1 (Figure 3a), COL12 (Figure 3b), COL6A3 (Figure 3c), COL7 (Figure 3d), and FBN2 (Figure 3f) were largely absent, whereas only sparse and irregular FBN1 (Figure 3e) fibers were detected. These findings are consistent with the secretion defects observed in the proteomic and biochemical analyses and demonstrate that selective export defects caused by loss of TANGO1 ultimately translate into profound alterations of ECM architecture. Interestingly, HSP47-deficient cells displayed a distinct phenotype characterized by the formation of unusually thick and irregular collagen fibers (Figure 3a-f). This observation is consistent with previous studies demonstrating that HSP47 regulates collagen folding and assembly rather than ER export ^10^. Thus, while both proteins influence ECM organization, they appear to do this through fundamentally different mechanisms.

**Figure 3.**
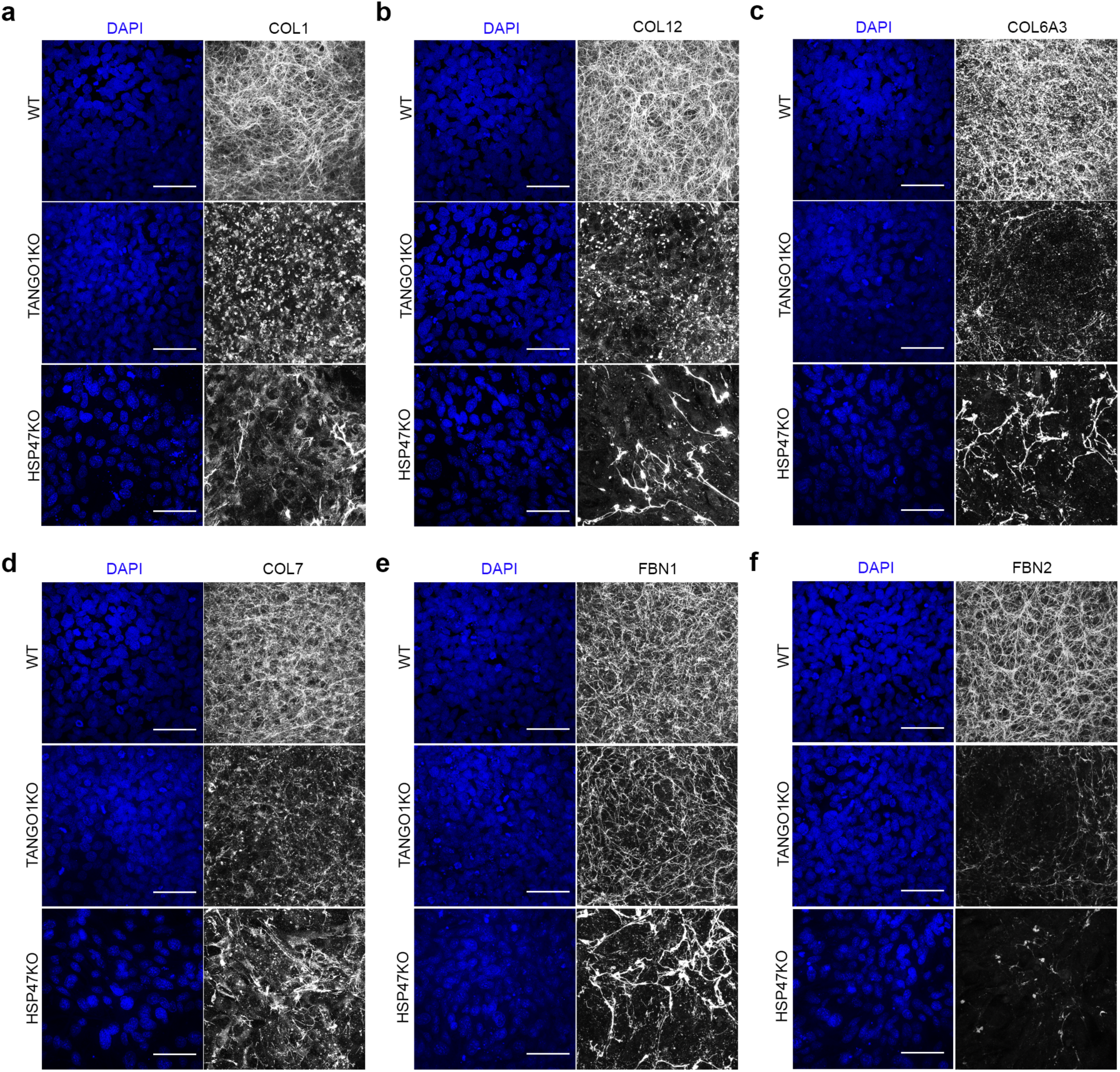
TANGO1-deficient cells exhibit ECM formation defects. **(a-f)** Immunofluorescence analysis of WT, TANGO1 KO and HSP47 KO cells cultured in the presence of ascorbic acid for 7 days, stained for type I collagen (COL1, a), type XII collagen (COL12, b), type VI collagen alpha 3 chain (COL6A3, c), type VII collagen (COL7, d), fibrillin-1 (FBN1, e) and fibrillin-2 (FBN2, f). Nuclei stained with DAPI. Scale bars, 50 μm. n = 3 independent experiments.

Since quantitative secretome analyses revealed distinctly different secretion phenotypes in TANGO1- and HSP47-deficient cells, we asked whether proteins that normally rely on TANGO1 for their secretion remain dependent on TANGO1 when HSP47 is absent (Ext. Data Figure 1). To address this question, we transiently depleted TANGO1 by siRNA in HSP47 wild-type and knockout cells. Consistent with our earlier observations, siRNA-mediated depletion of TANGO1 in wild-type cells resulted in reduced secretion and increased intracellular accumulation of COL7, COL12, and FBN2, whereas secretion of FBN1 remained largely unaffected (Ext. Data Figure 1a). Notably, a similar phenotype was observed when TANGO1 was depleted in HSP47 knockout cells. These results indicate that the export of these ECM proteins remains dependent on TANGO1 even in the absence of HSP47. We next examined whether these secretion defects affected ECM assembly. In wild-type cells, transient depletion of TANGO1 reduced the formation of fibrillar networks composed of COL1, COL12 and FBN2 two days after ascorbic acid addition, consistent with the phenotype observed in TANGO1 knockout cells. In HSP47 knockout cells, ECM assembly was compromised, and additional depletion of TANGO1 further reduced matrix formation, indicating that TANGO1 contributes to ECM formation also in the absence of HSP47 (Ext. Data Figure 1b-d).

### Loss of TANGO1-dependent export impairs ECM assembly and is rescued by TANGO1 re-expression

To directly assess the trafficking dynamics of TANGO1-dependent cargo, we performed synchronized secretion experiments using the RUSH (Retention Using Selective Hooks) system ^21^ (Ext. Data Figure 2a,b). In wild-type cells, SBP-tagged COL7 (Ext. Data Figure 2c) and FBN2 (Ext. Data Figure 2e) rapidly exited the ER following biotin addition and assembled into extracellular fibers. In contrast, both proteins remained strongly colocalized with the ER marker HSP47 in TANGO1 knockout cells and failed to form extracellular fibers, indicating impaired ER export. By comparison, trafficking of FBN1-SBP (Ext. Data Figure 2d) was largely unaffected by loss of TANGO1, as extracellular fibrillin fibers appeared with similar kinetics in both wild-type and knockout cells. These results demonstrate that TANGO1 controls ER export of selected ECM cargo proteins rather than global secretory pathway function.

Next, we reconstituted TANGO1 KO WI-26 cells by stably re-expressing HA-tagged TANGO1. Importantly, reconstitution of TANGO1 expression fully reversed the ECM secretion that was observed specifically for selected cargo such as COL1A2, COL12, COL6A3, COL7 and FBN2 in TANGO1 KO cells, showing that the ECM secretion defects are directly linked to the loss of TANGO1 (Ext. Data Figure 3a). Moreover, reconstituted TANGO1 KO cells also exhibited extracellular assembly of collagen (Ext. Data Figure 3b-d) and fibrillin fibers (Ext. Data Figure 3e), reversing the knockout phenotype. Confocal microscopy of ECM proteins in the TANGO1-deficient cells revealed a strong overlap of HSP47 with the retained COL12 (Ext. Data Figure 3f) and COL1 (Ext. Data Figure 3g), which was absent in control cells. Re-expression of TANGO1-HA in the TANGO1 KO cells led to a clearance of intracellularly accumulated COL12 and COL1, as well as HSP47, and the appearance of extracellular fibers. Altogether, these findings indicate that TANGO1 deficiency causes HSP47-independent ER retention of defined ECM cargo, rather than a general disruption of ERES function.

### Skeletal dysplasia-causing mutants in the MOTH domain of TANGO1 show selective ER export defects

To investigate whether disruption of TANGO1 cargo recognition contributes to human disease, we analyzed several previously unreported *MIA3* variants identified in patients presenting with skeletal dysplasia. Genetic analysis revealed six variants across four unrelated families, including missense variants affecting highly conserved residues within the luminal MOTH domain (Figure 4a) as well as truncating and splicing variants predicted to remove large parts of the luminal MOTH domain (Ext. Data Figure 4a-y). Based on the structure of the TANGO1 MOTH domain ^12^, these variants are expected to disrupt folding and to represent loss of function MOTH variants. A detailed description of patient recruitment, genetic analysis and clinical findings is provided in the Methods section and as Supplementary Information (Ext. Data Figure 4a-y).

**Figure 4.**
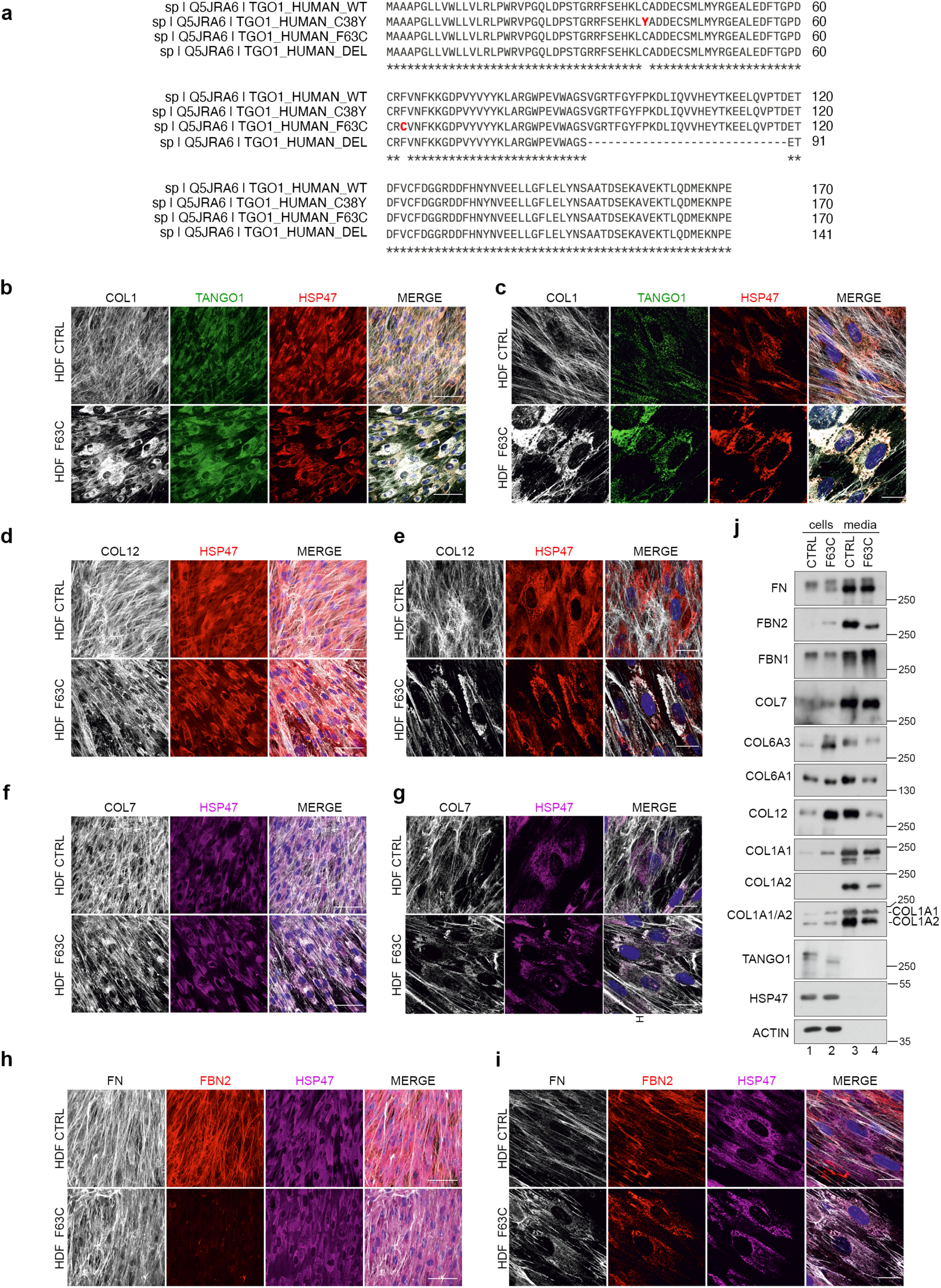
A single point mutation in the TANGO1 SH3L/MOTH domain blocks selective ER export of bulky ECM proteins. **(a)** Alignment of vertebrate TANGO1 SH3-like/MOTH domain amino acid sequence highlighting a high degree of conservation of the Cys38 and Phe63 residues. **(b-c)** Overview (z projection) (b) and high magnification (c) of immunofluorescence analysis of control and F63C HDFs, cultured in the presence of ascorbic acid for 7 days, stained for type I collagen (COL1), HSP47 and TANGO1. n = 3 independent experiments. **(d-e)** Overview (z projection) (d) and high magnification (e) of immunofluorescence analysis of control and F63C HDFs, cultured in the presence of ascorbic acid for 7 days, stained for type XII collagen (COL12) and HSP47. n = 3 independent experiments. **(f-g)** Overview (z projection) (f) and high magnification (g) of immunofluorescence analysis of control and F63C HDFs, cultured in the presence of ascorbic acid for 7 days, stained for type VII collagen (COL7) and HSP47. n = 3 independent experiments. **(h-i)** Overview (z projection) (h) and high magnification (i) of immunofluorescence analysis of control and F63C HDFs, cultured in the presence of ascorbic acid for 7 days, stained for fibronectin (FN), fibrillin-2 (FBN2) and HSP47. Nuclei stained with DAPI. Scale bars for overviews 50 μm and for high magnification 10 μm. n = 3 independent experiments. **(j)** Western blot analysis of cell lysates (cells) and supernatants (media) of ctrl and F63C HDFs, cultured for 24 h in the presence of ascorbic acid using the indicated antibodies. n = 3 independent experiments.

To assess the functional consequences of the two missense variants, as their effect on domain integrity is the least severe, we analyzed primary dermal fibroblasts derived from a patient carrying a p.Phe63Cys variant (hereafter F63C) in the TANGO1 MOTH domain. Immunofluorescence analysis revealed that patient fibroblasts, in contrast to control fibroblasts that formed an extensive extracellular fibrillar matrix, exhibited marked ER accumulation of several ECM proteins, including COL1 (Figure 4b,c), COL12 (Figure 4d,e), COL7 (Figure 4f,g) and FBN2 (Figure 4h,i), as revealed by colocalization with HSP47. Consistent with these observations, immunoblot analysis demonstrated a selective reduction in secretion of COL1A2, COL6A3, COL12 and FBN2, whereas secretion of COL1A1, COL6A1 and FBN1 was largely preserved (Figure 4j). These findings are in line with the cargo-selective secretion defects observed in TANGO1 knockout cells.

To further dissect the role of the MOTH domain, we reconstituted TANGO1 knockout cells with wild-type TANGO1 or patient-derived variants. Expression of wild-type TANGO1 restored secretion and extracellular assembly of TANGO1-dependent cargo proteins (Figure 5). In contrast, expression of the C38Y or F63C variants failed to rescue secretion defects, resulting in persistent intracellular retention of COL1 (Figure 5a,g), COL12 (Figure 5b,g), COL6A3 (Figure 5c,g), COL7 (Figure 5d,g) and FBN2 (Figure 5f,g). Intriguingly, FBN1 fibers were largely diminished in both cell lines expressing TANGO1 mutants, even though FBN1 secretion was largely unaffected, compared to control cells (Figure 5e,g). Importantly, both mutants localized to SEC31-positive ER exit sites, comparably to wild-type TANGO1, indicating that the observed secretion defects do not stem from impaired ERES recruitment (Ext. Data Figure 5a,b). In contrast, despite their ERES localization, both mutants showed a low degree of colocalization with the TANGO1 cargo protein COL12, suggesting impaired cargo recruitment at these locations (Ext. Data Figure 5c,d).

**Figure 5:**
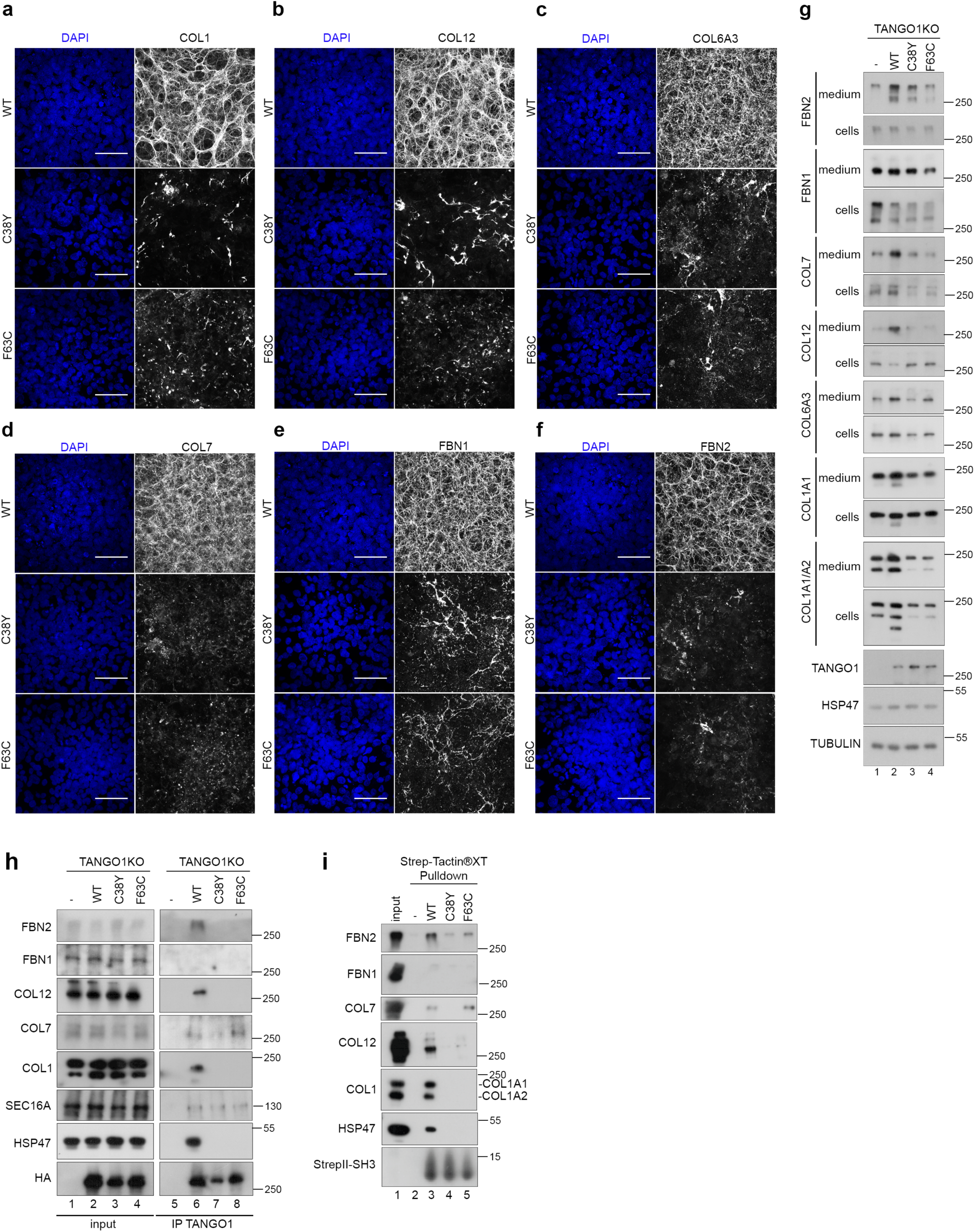
TANGO1 patient mutants fail to bind to ECM cargo proteins and do not rescue ER export of bulky cargo. **(a-f)** Immunofluorescence analysis (z projections) of TANGO1 KO cells reconstituted with TANGO1-HA WT, C38Y and F63C mutants, cultured in the presence of ascorbic acid for 7 days, stained for type I collagen (COL1, a), type XII collagen (COL7, b), type VI collagen alpha 3 chain (COL6A3, c), type VII collagen (COL7, d), fibrillin-1 (FBN1, e) and fibrillin-2 (FBN2, f). Nuclei stained with DAPI. Scale bars, 50 μm. n = 3 independent experiments. **(g)** Western blot analysis of cell lysates (cells) and supernatants (media) of TANGO1 KO cells and TANGO1 KO cells stably reconstituted with TANGO1-HA WT, C38Y and F63C mutants, cultured for 24 h in the presence of ascorbic acid using the indicated antibodies. n = 3 independent experiments. **(h)** TANGO1 Immunoprecipitation analysis using lysates of TANGO1 KO cells and TANGO1 KO cells stably reconstituted with TANGO1-HA WT, C38Y and F63C mutants, cultured for 24 h in the presence of ascorbic acid, followed by western blot analysis using the indicated antibodies. n = 3 independent experiments. **(i)** Strep-TactinXT pulldown assay using recombinantly expressed TwinStrep-tagged TANGO1-SH3 WT, C38Y and F63C mutants, incubated with lysate of WI26-SV40 cells, cultured for 24 h in the presence of ascorbic acid, followed by western blot analysis using the indicated antibodies. n = 3 independent experiments.

Given that the disease-associated mutations reside within the predicted cargo-binding domain of TANGO1, we next examined whether these variants impair interactions with ECM cargo proteins. Co-immunoprecipitation experiments using TANGO1 knockout cells reconstituted with wild-type or the disease-associated TANGO1 variants, revealed that wild-type TANGO1 interacts with several ECM proteins identified as TANGO1-dependent cargo, including COL1, COL7, COL12 and FBN2, but not FBN1 (Figure 5h). In contrast, the C38Y mutant failed to interact with these proteins, while the F63C variant displayed strongly reduced binding (Figure 5h). Consistent with these results, affinity pulldown assays using recombinant MOTH domains demonstrated direct binding of the wild-type domain to COL1, COL7, COL12 and FBN2, but not to FBN1 (Figure 5i). Notably, the C38Y variant completely abolished cargo binding, while the F63C variant showed only weak residual interactions. These findings indicate that the MOTH domain mediates selective recognition of ECM cargo proteins and that disease-associated mutations disrupt these receptor-cargo interactions.

### TANGO1 directly recognizes specific ECM chains through a highly conserved tyrosine residue in its MOTH domain

To further characterize the molecular basis of cargo selectivity, we performed surface plasmon resonance (SPR) binding assays using purified type VI collagen chains and recombinant MOTH domain of TANGO1 (Figure 6a). In line with our findings from cell culture experiments, no binding was detected for COL6A1 (Figure 6b) or COL6A2 (Figure 6c), while the COL6A3 chain (Figure 6d) specifically interacted with the immobilized TANGO1 MOTH domain. Mapping experiments employing truncation variants revealed that binding is mediated by the C3-C4 domains within the C-terminal non-collagenous region of COL6A3, but not the individual C2 and C5 domains (Figure 6e-i). Similarly, binding assays with fibrillin fragments demonstrated selective interaction between TANGO1 and an N-terminal fragment of FBN2 (rF86), whereas no binding was observed for other FBN2 fragments or the respective corresponding FBN1 fragments (Ext. Data Figure 6a-i). These findings demonstrate that TANGO1 directly recognizes specific ECM cargo chains rather than entire collagen molecules or fibrillin proteins.

**Figure 6.**
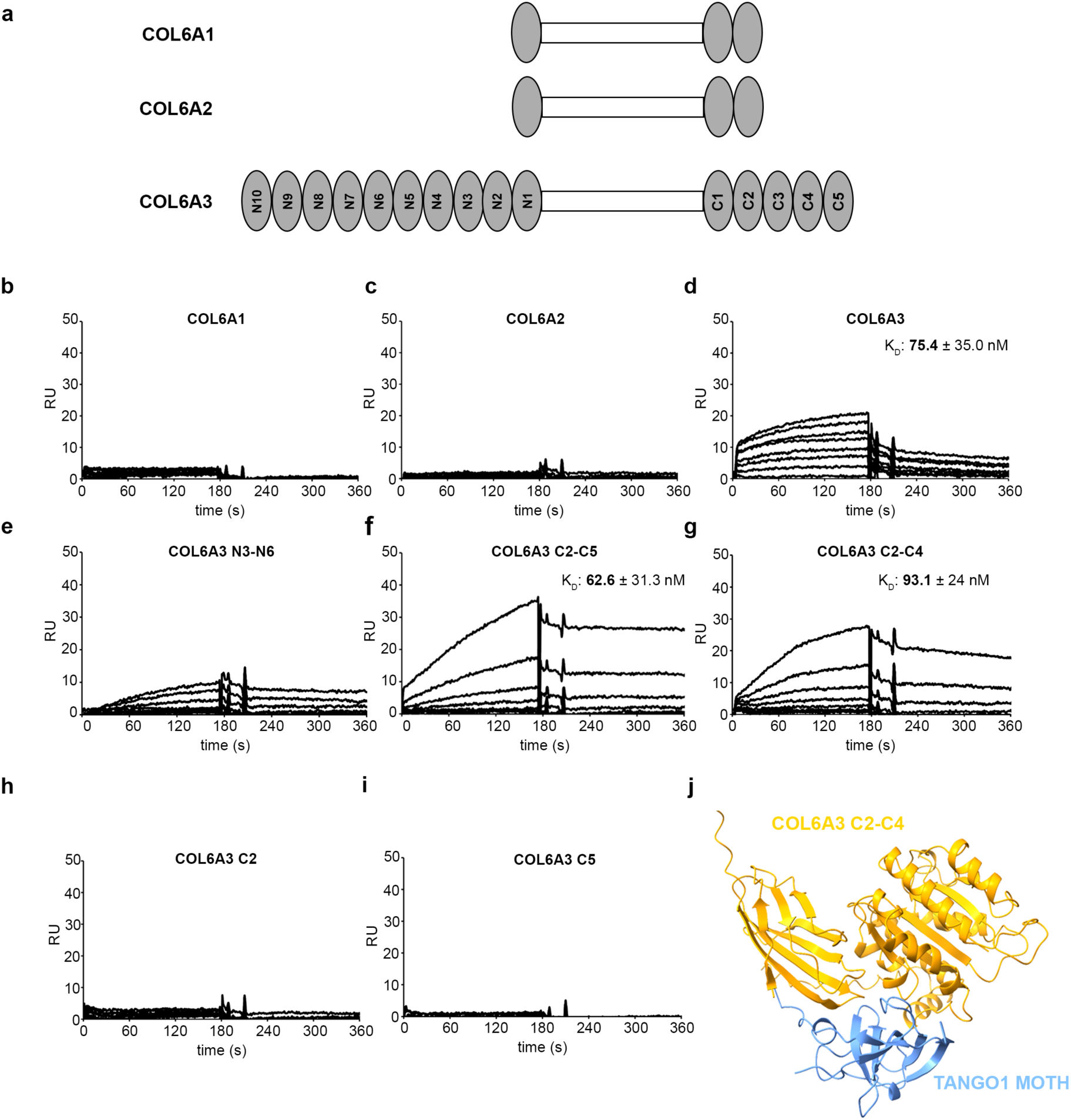
TANGO1 selectively binds to the C3-C4 domains of COL6A3. **(a)** Domain structures of human type VI collagen chains (COL6A1, COL6A2 and COL6A3). **(b-i)** SPR sensorgrams obtained either of the interaction between TANGO1 MOTH domain (immobilized on the sensor chips in similar quantities) and COL6A1 (b), COL6A2 (c), COL6A3 (d) or the respective COL6A3 fragments (e-i) (flown over as soluble ligands) in a dilution series from 0 to 320 nM. Ligand binding to the immobilized proteins on the chip is shown as response units on the y-axis (n=3). **(j)** Alphafold3 Model of the complex between the TANGO1 MOTH domain and COL6A3 C2-C4 domains.

Structural modelling of the interaction between the TANGO1 MOTH domain and the COL6A3 C3-C4 region suggested that several residues of the TANGO1 luminal domain contribute to the cargo-binding interface (Figure 6j and Ext. Data Figure 7a-c). The predicted interaction interface is highly conserved and predominantly exposes tyrosine on position 75 of TANGO1. This residue is invariant across vertebrate TANGO1 orthologs (Figure 7a,b), suggesting an evolutionarily conserved role in cargo recognition. Indeed, mutation of Tyr75 to arginine abolished binding to COL6A3 and its C-terminal fragments as shown by SPR measurements (Ext. Data Figure 7d,e). To assess the functional relevance of this residue, we generated a full-length TANGO1 Y75R mutant and stably reconstituted TANGO1 knockout cells with either wild-type or mutant TANGO1. Immunofluorescence analysis showed that re-expression of wild-type TANGO1 restored extracellular fibrillar networks of multiple cargo proteins, including COL1 (Figure 7c), COL12 (Figure 7d), COL6A3 (Figure 7e), COL7 (Figure 7f), and FBN2 (Figure 7g). By contrast, cells expressing the Y75R TANGO1 mutant displayed a strong reduction in extracellular deposition of these proteins, with staining appearing predominantly intracellular, consistent with a secretion defect (Figure 7c-g). As expected, FBN1, which we demonstrated is secreted independently of TANGO1, showed a diffuse extracellular fibrillar pattern (Figure 7h). Immunoblot analysis of cell lysates and conditioned media corroborated these findings (Figure 7i). Wild-type TANGO1 restored secretion of COL1A2, COL6A3, COL12, COL7, and FBN2, whereas expression of the Y75R mutant failed to rescue their export, leading to intracellular accumulation. Secretion of FBN1 and COL6A1 was comparatively less affected (Figure 7i), consistent with their reduced TANGO1 dependency observed in both the secretome analysis and in patient fibroblasts. Together, these results demonstrate that Tyr75 within the MOTH domain is required for selective recognition and ER export of specific ECM cargo chains, and that its mutation disrupts cargo binding and phenocopies the secretion defects of TANGO1 knockout cells.

**Figure 7.**
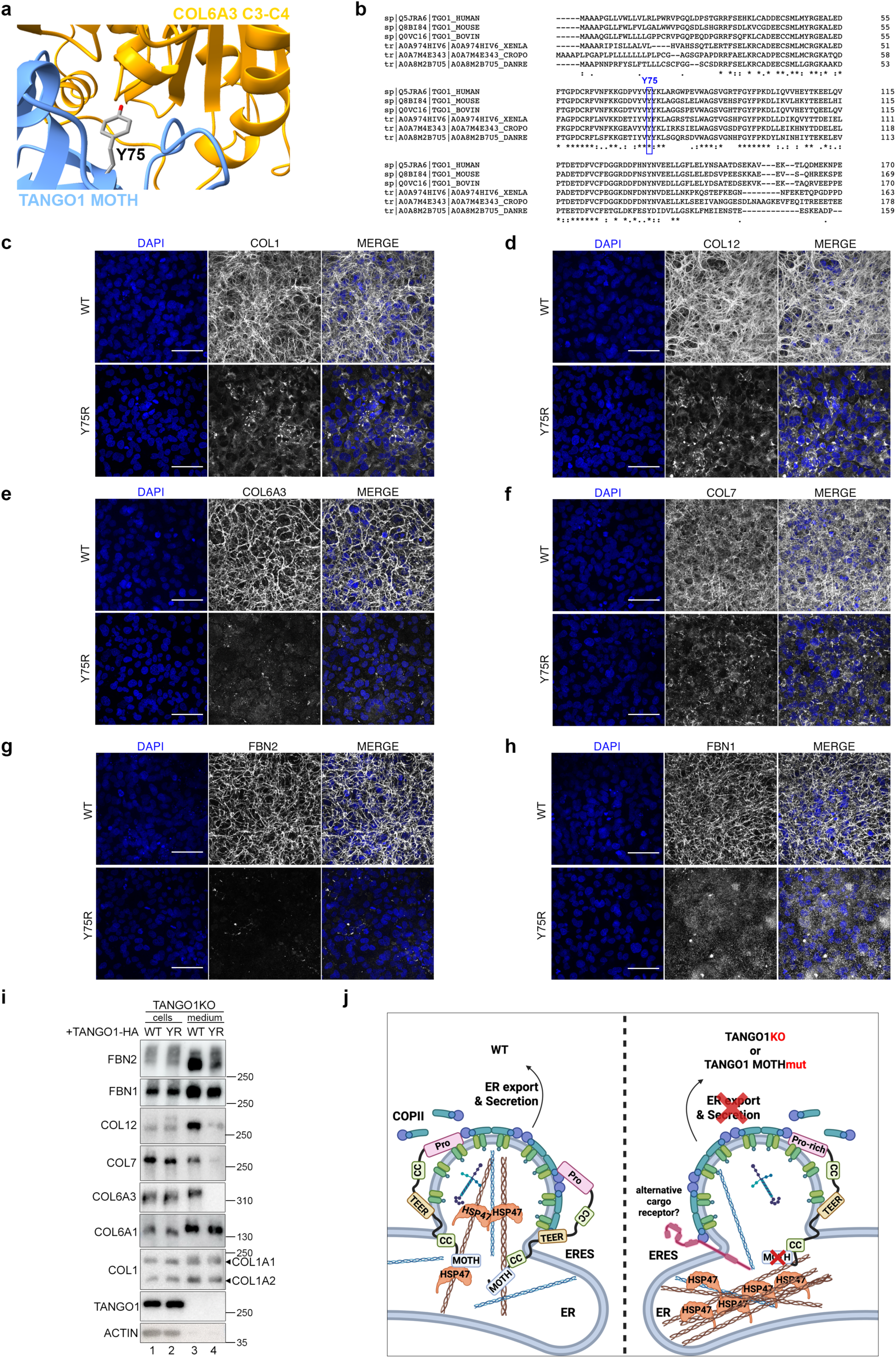
TANGO1 Y75R mutation diminishes bulky cargo export from ERES. **(a)** Close-up of interaction site of TANGO MOTH domain with COL6A3 C3-C4 domains with the key interacting residue Tyr75 shown in stick representation. **(b)** The human TANGO1 Tyr75 is highly conserved among vertebrates. **(c-h)** Immunofluorescence analysis (z projections) of TANGO1 KO cells reconstituted with TANGO1-HA WT and Y75R mutant, cultured in the presence of ascorbic acid for 7 days, stained for type I collagen (COL1, c), type XII collagen (COL12, d), type VI collagen alpha 3 chain (COL6A3, e), type VII collagen (COL7, f), fibrillin-2 (FBN2, g) and fibrillin-1 (FBN1, h). Nuclei stained with DAPI. Scale bars, 50 μm. n = 3 independent experiments. **(i)** Representative western blot analysis of cell lysates (cells) and supernatants (media) of TANGO1 KO cells stably reconstituted with TANGO1-HA WT and Y75R mutant, cultured for 24 h in the presence of ascorbic acid using the indicated antibodies. n = 3 independent experiments. **(j)** Schematic working model of TANGO1 selectively binding to bulky ECM cargo at ERES and how loss of TANGO1 or mutation in the cargo-binding MOTH domain leads to ER retention of ECM proteins. See text for more details.

Based on these findings, we propose a model in which TANGO1 functions as a selective ECM cargo receptor at ER exit sites rather than a general bulky cargo export factor (Figure 7j). In wild-type cells, the luminal MOTH domain directly engages specific ECM cargo chains and promotes their export through TANGO1-organized ERES. In the absence of TANGO1, or upon disruption of the cargo-binding interface, these proteins accumulate in the ER while other ECM cargoes may exit cells via putative alternative pathways.

## Discussion

Our study demonstrates that TANGO1 functions as a selective ECM cargo receptor rather than a general facilitator of bulky cargo secretion. We show that TANGO1 directly recognizes specific ECM proteins through its luminal MOTH domain, independently of HSP47, and that disruption of this interaction underlies TANGO1-associated skeletal dysplasia. Together, these findings identify cargo selectivity as a fundamental principle of TANGO1 function and provide a mechanistic framework for ECM cargo recognition and export at ER exit sites.

### TANGO1 and HSP47 Play Unique but Complementary Roles in ECM Homeostasis

Efficient ECM secretion requires the coordinated action of folding chaperones and specialized export machinery at ERES, yet the principles governing TANGO1 cargo selectivity and its relationship with the collagen-specific chaperone HSP47 have remained unresolved ^4,5,8,11^. Comparing the secretomes of TANGO1- and HSP47-deficient cells, resolves these as mechanistically distinct activities. TANGO1 is required for the export of a defined subset of ECM cargo including COL6A3, COL1A2, COL12, COL7 and FBN2, while leaving size-matched proteins such as COL1A1, COL6A1 and FBN1 unaffected, establishing that cargo size alone does not dictate TANGO1 dependency, and TANGO1 is not a general secretion factor ^22^ but instead selectively facilitates the secretion of specific ECM proteins. Notably, matrix assembly was also impaired for cargoes whose secretion was largely TANGO1-independent, such as collagen I and FBN1. For collagen I this may reflect chain-specific dependency, as COL1A2 but not COL1A1 secretion was reduced. In fact, COL1A1 homotrimers are known to alter matrix properties and accompany fibrotic disease ^23,24^. More broadly, the concurrent loss of collagen-modifying enzymes from the TANGO1-dependent secretome ^17–19^, offers a parsimonious explanation for why matrix organization can fail even when a cargo is itself secreted normally. TANGO1 thus shapes the ECM both directly, through cargo export, and indirectly, by controlling the secretion of ECM maturation factors.

Consistent with its established role in stabilizing the collagen triple helix ^8,11,25^, HSP47 acts within the ER lumen at the folding step rather than in bulky cargo export. Critically, many TANGO1-dependent cargoes are still secreted in the absence of HSP47, showing that cargo recognition by TANGO1 does not require this chaperone. Together, these data place HSP47 and TANGO1 to separable steps in collagen biogenesis, namely folding and selective export, respectively, and are incompatible with models in which HSP47 serves as the adaptor that links collagen cargo to TANGO1.

### TANGO1 selectively binds bulky cargo proteins via a conserved motif

The defining advance of this study is that TANGO1 recognizes cargo directly, rather than indirectly through HSP47 or solely as a membrane scaffold coordinating ERES. The MOTH domain discriminates among closely related chains, for instance binding COL6A3 but not COL6A1 or COL6A2, and FBN2 but not FBN1, through an invariant, evolutionarily conserved tyrosine within a surface pocket whose mutation abolishes binding and recapitulates the secretion defect in full TANGO1 knockouts. Biologically, this establishes sequence- or structure-encoded features in the cargo, not size, as the determinant of TANGO1 dependency, and defines TANGO1 as a bona fide cargo receptor. Selective engagement of COL6A3 further raises the possibility that TANGO1 participates in collagen VI biogenesis itself, coupling recognition of a single obligate subunit to the coordinated export of the assembled tetramer ^3^, extending TANGO1’s role from controlling which proteins are exported to influencing the assembly state of the matrix. That some large ECM proteins are still secreted without TANGO1 implies additional cargo receptors or export routes, whose identification will be important for understanding how cells maintain ECM homeostasis.

### Skeletal dysplasia-causing variants in TANGO1/MIA3 affect selective recognition of ECM cargo proteins

Variants in *MIA3* cause skeletal dysplasia characterized by impaired bone growth and connective tissue abnormalities ^13–15^, but the underlying mechanism has been unclear to date. Here we show that disease-associated missense mutations in the TANGO1 MOTH domain (C38Y and F63C) phenocopy the selective secretion defects of TANGO1 knockout cells while retaining ER exit site localization and interactions with the secretory machinery, therefore preserving the structural, ERES-organizing role of TANGO1. This suggests that impaired folding of the TANGO1 MOTH domain disrupts interaction with specific ECM cargo, causing their ER accumulation and failure to be secreted and incorporated into the ECM. Disease thus arises not from a general collapse of bulky cargo export, but from the failure of selective cargo recognition, reinforcing that this step, rather than TANGO1’s scaffolding function, is the physiological role of TANGO1 in cells.

This framework also resolves an apparent paradox. Monogenic loss of individual TANGO1-dependent cargoes does not reproduce the skeletal phenotype observed in patients carrying TANGO1 loss of function variants: *COL12A1* variants cause a myopathy-related connective tissue disorder with joint laxity ^26,27^, and *COL6A3* variants cause Bethlem myopathy or Ullrich congenital muscular dystrophy ^28,29^; neither a primary skeletal dysplasia. This is most readily explained by the combinatorial nature of the TANGO1-dependent defect: TANGO1 loss simultaneously impairs export of multiple ECM components (COL1A2, COL6A3, COL7, COL12, FBN2) and collagen-modifying enzymes (LOXL2, prolyl hydroxylases), and it is this concurrent depletion, rather than loss of any single component, that likely determines phenotype severity. The retention of COL6A3 has specific consequences for collagen VI assembly, which proceeds intracellularly through antiparallel dimers and tetramers in which COL6A3 plays a non-redundant structural role ^30,31^, so ER-trapped COL6A3 would be expected to block export of assembled tetramers altogether. This is consistent with our finding in TANGO1-deficient or cells expressing TANGO1 MOTH domain mutants, that COL6A3 is retained while COL6A1/COL6A2 secretion is relatively preserved, presumably as unassembled chains exported via alternative secretory routes. Whether these chains contribute functionally to matrix organization is unclear, but Col6a3-null mice, which show skeletal and muscle phenotypes, indicate that COL6A3 loss profoundly disrupts pericellular matrix architecture in collagen-rich tissues ^30,31^. Similarly, mutations in SEC23A, a COPII coat component, cause cranio-lenticulo-sutural dysplasia through simultaneous impaired export of multiple collagens and matrix proteins ^32^. Together, the findings presented here identify defective cargo selectivity, rather than global secretion failure, as the mechanism underlying TANGO1-associated skeletal dysplasia.

## Methods

### Cell culture and treatments

All cell lines were grown at 37°C, 5% CO_2_. Human male diploid lung WI-26 SV40 fibroblasts (here termed WI-26 cells; #CCL-95.1, ATCC; RRID: CVCL_2758) were cultured in DMEM/F12 GlutaMAX medium (#31331093, Gibco) containing 10% FBS. Normal (non-transformed) human male foreskin fibroblasts HFF-1 (HFFs; #SCRC-1041, ATCC; RRID: CVCL_3285), human female embryonic kidney HEK293FT cells (#R70007, Invitrogen; RRID: CVCL_6911) and human patient-derived dermal fibroblasts were cultured in high-glucose DMEM + L-Glutamine (#61965026, Gibco), containing 10% FBS. All media were supplemented with Penicillin-Streptomycin (#15140122, Gibco; #P4333-100ML, Sigma).

The identities of the HEK293FT and WI-26 cells were validated using the Short Tandem Repeat (STR) profiling service, provided by Multiplexion GmbH. No commonly misidentified cell lines were used in this study. All cell lines were regularly tested for *Mycoplasma* contamination, using a PCR-based approach and were confirmed to be *Mycoplasma*-free.

For matrix production experiments, cells were cultured for the indicated time points in the presence of 150 μg/ml ascorbic acid-2-phosphate (A8960, Sigma Aldrich).

### Patients and genetic analysis

Six previously unreported patients and three fetuses from four families of different ancestries (Italian, Iranian, Spanish and Slovenian) were included in this study after obtaining written informed consent. Clinical data were collected for each affected subject by the clinicians from the participating centers using a standardized questionnaire. The available radiological imaging was reviewed by a radiologist expert in skeletal dysplasia (A.C.O.), and orthopantomograms were reviewed by an expert in hereditary dental disorders (A.B.-Z.). The ethics committee of Instituto Giannina Gaslini (Genoa - Italy) and the other participating centres gave ethical approval for this work.

Exome sequencing was performed on DNA isolated from peripheral blood of affected patients or from amniocytes for fetuses and their parents when available. Slightly different analysis platforms were used in the respective collaborating centers according to the Burrows-Wheeler Aligner (BWA)/The Genome Analysis Tool kit (GATK)-based pipeline. Sanger sequencing with standard methods was performed for candidate variants’ validation and familial segregation. All *MIA3* variants are reported according to the transcript NM_198551.4 and classified according to the American College of Medical Genetics and Genomics (ACMG) and the Association for Molecular Pathology (AMP) variant classification system.

### Isolation of patient fibroblasts

Human dermal fibroblasts (Fibroblasts) were isolated from skin biopsies as previously described ^33^. The TANGO1 F63C expressing cells cells were collected from patient II:2 of Family 1. Fibroblast cultures were maintained in Dulbecco’s Modified Eagle Medium (DMEM) supplemented with 10% fetal bovine serum (FBS, Hyclone), and 1% Pen-Streptomycin at 37 °C with 5% CO_2_.

### Antibodies

A list of all primary antibodies used in this study is provided in Supplementary Table 3.

### Plasmids and molecular cloning

Plasmid expression vectors were generated by cloning PCR-amplified cDNAs, using appropriate primers (Supplementary Table 4). cDNA was generated from total RNA isolated from human WI-26 fibroblasts using a standard Trizol/chloroform-based extraction (15596018, Thermo Fisher Scientific) using Superscript II (#18064014, Thermo Fisher Scientific). For the construction of the TANGO1-HA expression vector pITR-TTP-hTANGO1-HA WT, C38Y, F63C and Y75R, human TANGO1 (NM_198551.4) was amplified from WI-26 cDNA and cloned into the SfiI/NotI restriction sites of the pITR-TTP plasmid ^16^ using the NEB HiFi Builder (#E2621, NEB). The RUSH construct for COL7A1 (pIRES-Str-KDEL-hCOL7-SBP) was generated by amplification of human COL7A1 (NM_000094.4) from pcDNA-FLAG-hCOL7A1 (kind gift from Prof. Alexander Nyström, University of Freiburg) and cloning of the cDNA into the AscI and FseI restriction sites of the Str-KDEL_ManII-SBP-EGFP vector (Addgene plasmid #65252) ^21^. Next, the SBP (Streptavidin-Binding Peptide) tag was amplified from Str-KDEL_ManII-SBP-EGFP and inserted in-frame with COL7A1 using the FseI and PacI restriction sites. The respective RUSH constructs for FBN1 (pIRES-Str-KDEL-hFBN1-SBP) and FBN2 (pIRES-Str-KDEL-hFBN2-SBP) were generated by amplification of human FBN1 (NM_000138.5) and FBN2 (NM_001999.4) from WI-26 cDNA and assembled with the PCR amplified SBP tag sequence from Str-KDEL_ManII-SBP-EGFP using the NEB HiFi Builder and cloning into the AscI and PacI restriction sites of the Str-KDEL_ManII-SBP-EGFP vector.

TwinStrep-tagged TANGO1 MOTH domain WT, C38Y, F63C (TST-MOTH WT, C38Y and F63C) were PCR amplified from pITR-TTP-hTANGO1-HA WT, C38Y or F63C and subcloned into the NheI/NotI restriction sites of the pITR-TTP ^16^ in frame with the BM40 signal peptide and the TwinStrep-tag. The TwinStrep-tagged TANGO1 MOTH Y75R mutant (TST-MOTH Y75R) was generated by quick change site-directed mutagenesis using the pITR-TTP-TST-SH3L/MOTH WT construct as template.

The pSpCas9^34^-2A-Puro (PX459) V2.0 plasmid ^35^. (Addgene plasmid #62988) was used for the generation of TANGO1 and HSP47-deficient WI-26 cells. In brief, double-stranded DNA oligos that encode guide RNAs (gRNAs) against target genes were cloned into the BbsI restriction sites of the PX459 vector. The oligo sequences used for the sgRNA expression plasmids to generate the TANGO1 and HSP47 KO lines are provided in Supplementary Table 4.

The generation of expression constructs for the recombinant fibrillin-1 and fibrillin-2 fragments were described previously ^36,37^. The generation of the murine Col6A3 C2, C2-C5, C5 and N3-N6 were described previously ^38–41^. For the expression constructs for murine Col6a1, murine Col6a2, murine Col6a3, cDNA of E19 embryos was used and cloned into the pITR-TTP vector. For the murine Col6A3 C2-C4 fragment, the published murine Col6A3 C2-C5 was used as a template ^38^ (Supplementary Table 4).

All restriction enzymes were purchased from Thermo Fisher. The integrity of all constructs was verified by sequencing.

### Plasmid DNA transfections

Plasmid DNA transfections were performed using the X-tremeGENE HP DNA transfection reagent (#06366236001, Roche) in a 2:1 DNA/transfection reagent ratio when the WI-26 cells reached approximately 70% confluency, according to the manufacturer’s instructions. Twenty-four hours post-transfection, cells were either lysed for immunoblotting or fixed for immunofluorescence.

### Gene silencing experiments

Transient knockdown of HSP47/SERPINH1 was performed using siGENOME (pool of four) gene-specific short interfering RNAs (siRNAs) (Horizon Discoveries). An siRNA duplex targeting the Renilla reniformis luciferase gene (RLuc) (#P-002070-01-50, Horizon Discoveries) was used as control. Transfections were performed using 20 nM siRNA and the HiPerfect transfection reagent (#301705, QIAGEN)), according to the manufacturer’s instructions. Cells were collected or fixed 96 h post transfection, and knockdown efficiency was verified by immunoblotting.

### Generation of knockout cell lines

The TANGO1 and HSP47 knockout cell lines were generated using the PX459-based CRISPR/Cas9 method, as described elsewhere ^35^. In brief, WI-26 cells were transfected with sgRNA-expressing vectors and were selected 36 hours post-transfection with puromycin (2 μg/ml) (#A1113803, Gibco) for 5 days. Single cell clones were picked using cloning cylinders (#CLS31668, Sigma-Aldrich) and knockout clones were validated by genomic DNA sequencing and immunoblotting. An empty PX459 vector was used to generate matching control cell lines.

### Generation of stable cell lines

Reconstituted TANGO1 KO WI-26 cell line, stably expressing TANGO1-HA WT, C38Y or F63C were generated by reconstituting TANGO1 KO WI-26 cells using a doxycycline-inducible sleeping-beauty-based transposon system as described before ^16^. In brief, cells were transfected with the transposon-flanked pITR-TTP-TANGO1-HA plasmids (WT, C38Y, F63C and Y75R TANGO1) described above, together with the transposase expressing pCMV-Trp vector. At 24 h post transfection, puromycin (2 μg/ml) was added to the medium and cells were selected for 5 days. Single-cell colonies were picked using cloning cylinders (#CLS31668, Sigma-Aldrich) and expanded. Clones that express similar levels of WT, C38Y or F63C TANGO1 were used in follow-up experiments.

HEK293FT stably expressing TwinStrep-tagged TANGO1 MOTH domain WT, C38Y, F63C and Y75R were generated by using a doxycycline-inducible sleeping-beauty-based transposon system as described above.

### Cell lysis, preparation of supernatant samples, and immunoblotting

For standard SDS-PAGE and immunoblotting experiments, cells from one well of a 6-well plate, at approximately 90% confluence, were lysed in-well with 300 μl ice-cold Triton lysis buffer (50 mM Tris-HCl pH 7.5, 0.5% Triton X-100, 150 mM NaCl, 0.1% SDS), supplemented with 1x EDTA-free cOmplete protease inhibitors (#11873580001, Roche) and 1x PhosSTOP phosphatase inhibitors (#4906837001, Roche). Lysates were cleared by centrifugation (12,000 x g, 15 min, 4°C) to remove debris and supernatants transferred to a new tube. Samples were boiled in 1x SDS sample buffer for 5 min at 95 °C (6x SDS sample buffer: 350 mM Tris-HCl pH 6.8, 30% glycerol, 600 mM DTT, 12.8% SDS, 0.12% bromophenol blue) or in SDS sample buffer without DTT (for FBN1 and FBN2 immunoblots).

To determine levels of secreted proteins, serum-free supernatants were centrifuged (2000 x g, 5 min, 4°C) to remove dead cells and debris. Cleared culture supernatants were concentrated using 3 kDa cut-off concentrator tubes (#516-0227P, VWR) according to the manufacturer’s instructions, and Laemmli loading buffer (1x final concentration; 6x SDS sample buffer: 350 mM Tris-HCl pH 6.8, 30% glycerol, 600 mM DTT, 12.8% SDS, 0.12% bromophenol blue) or Laemmli sample buffer without DTT (for FBN1 and FBN2 immunoblots) was added to the concentrated supernatants.

Protein samples were subjected to electrophoretic separation on SDS-PAGE and analyzed by standard Western blotting techniques. In brief, proteins were transferred to PVDF membranes (IPVH00010, Millipore). Membranes were blocked with 5% skim milk powder (#42590, Serva) in TBS-T buffer [50 mM Tris-HCl pH 7.4, 150 mM NaCl, 0.1% Tween-20 (#A1389, AppliChem)] and incubated with primary antibodies diluted in TBS-T or in 5% skim milk/TBS-T (for the goat anti-COL1 antibody), for 1 hour at room temperature or overnight at 4°C, followed by incubation with appropriate HRP-conjugated secondary antibodies in TBS-T or in 5% skim milk/TBS-T (for the anti-goat-HRP secondary antibody) for 1 hour at room temperature. Signals were detected by enhanced chemiluminescence (ECL) and immunoblot images were captured on films (#28906835, GE Healthcare; #4741019289, Fujifilm). Quantification of immunoblots was performed by densitometric analysis of the band intensities, using the Gel analysis tool of the ImageJ software ^42^. A list of all primary and secondary antibodies used in this study is provided in Supplementary Table 3.

### Co-immunoprecipitation (co-IP)

For co-IP experiments, TANGO1 KO cells stably expressing HA-tagged wild-type or C38Y or F63C mutant TANGO1, or empty vector control cells (1× 10 cm dish per condition) were cultured in the presence of 150 μg/ml ascorbic acid-2-phosphate. Cells were lysed 24 hours post-transfection in 500 μl Triton lysis buffer per dish (50 mM Tris-HCl pH 7.5, 1% Triton X-100, 150 mM NaCl, 1x EDTA-free cOmplete protease inhibitors, 1x PhosSTOP phosphatase inhibitors) using a glass Dounce homogenizer and incubated for 30 minutes on ice. Samples were cleared by centrifugation (12,000 x g, 10 min, 4°C) and incubated with 2 μg of anti-TANGO1 polyclonal antibody for 3-4 hours at 4 °C under constant agitation. Complexes were incubated overnight with 40 μl protein A agarose beads, washed 4x with lysis buffer, boiled in 1x Laemmli sample buffer (5 min, 95 °C), and separated by SDS-PAGE. Input samples (50 μl) were collected before pre-clearing, SDS sample buffer (1x final concentration) was added to the lysates, samples were boiled and analyzed by immunoblotting.

### Immunofluorescence and confocal microscopy

Immunofluorescence/confocal microscopy experiments were performed as described previously ^16^ with minor modifications. In brief, cells were grown on glass coverslips and treated as described in the figure legends. Samples were fixed for 10 min at room temperature with 4% formaldehyde (#252549, Sigma-Aldrich) in PBS or for 5 min at –20°C with 100% ice-cold methanol, permeabilized with 0.5% NP-40 (I8896, Sigma-Aldrich) in PBS for 10 min, blocked with 1% FBS (FBS.HP.0500, Bio&SELL) in PBS for 30 min, incubated with primary antibodies diluted in blocking buffer (1% FBS in PBS) for 1 hour at room temperature, washed 3x with blocking buffer, and incubated with appropriate highly cross-adsorbed secondary antibodies conjugated to Alexa Fluor 488, 555 or 647 (Thermo Fisher Scientific) for 1 hour at room temperature. Nuclei were stained with 0.1 μg/ml DAPI (#D9542, Sigma-Aldrich). Samples were mounted with Fluoromount-G mounting medium (#00-4958-02, Invitrogen). All images were acquired on an SP8 Leica confocal microscope (TCS SP8 X or TCS SP8 DLS, Leica Microsystems) using 63x or 100x objective lenses. Image acquisition was performed using the LAS X software (Leica Microsystems). Brightness and contrast were adjusted for visualization purposes using Fiji (https://imagej.net/software/fiji/downloads) ^43^. Alterations were applied to the entire image, keeping the parameters identical between all images of the same channel in each panel.

### Quantification of colocalization

Colocalization analysis in confocal microscopy experiments was performed as described before ^44,45^, using the Coloc2 plugin of the Fiji software ^43^. At least 50 individual cells from 5 independent representative images per condition per replicate (3-4 replicates per experiment) were used to calculate Manders’ colocalization coefficient ^22^ with automatic Costes thresholding ^34,46,47^. Outlines of individual cells were traced, excluding the area corresponding to the cell nucleus, to generate the region of interest (ROI) used for calculating the MCC to prevent false-positive colocalization due to automatic signal adjustments. MCC is defined as a part of the signal of interest (TANGO1-HA), which overlaps with a second signal (SEC31, COL12).

### Retention Using Selective Hooks (RUSH) assay

Synchronized trafficking of proteins in the secretory pathway ^21^ was achieved by transfecting WT and TANGO1 KO WI-26 cells with the Str-KDEL-containing COL7-SBP, FBN1-SBP or FBN2-SBP reporter constructs (see also Plasmids). Twenty-four hours post-transfection, cells were treated with 40 μM biotin (#Β4639, Sigma-Aldrich) for different time points as indicated in the figure legends, fixed and subjected to indirect immunofluorescence.

### Identification of *MIA3* variants

Exome sequencing revealed six novel *MIA3* variants in affected individuals as follows: compound heterozygous variant c.188T>G p.(Phe63Cys) and c.4654del p.(Leu1552Cysfs*15) in Family 1, a splicing homozygous variant c.354+2T>G in Family 2, compound heterozygous variant c.113G>A p.(Cys38Tyr) and c.3286_3287ins p.(Ser1096Phefs*54) in Family 3 and a nonsense homozygous variant c.2768T>G p.(Leu923*) in Family 4. Sanger sequencing confirmed segregation of the variants with the phenotype within these families. All variants were classified according to the ACMG/AMP criteria and were absent from the gnomAD database (https://gnomad.broadinstitute.org/) except for the c.2768T>G p.(Leu923*) variant that has been reported in the heterozygous state at very low frequency (1.86e-6). None of the variants were reported in a homozygous state in the gnomAD database. All nonsense and frameshift variants are predicted to result in a premature truncation of the transcript, likely leading to nonsense-mediated mRNA decay. The canonical variant c.354+2T>G is predicted to severely impair the protein structure through aberrant mRNA splicing. The missense variants affect highly conserved residues (Figure 4a). No other pathogenic/likely pathogenic variants were identified in the currently known skeletal dysplasia–related genes and no other candidate genes were identified in the affected individuals.

### Phenotypic spectrum associated with *MIA3* variants

The present *MIA3* cohort include six individuals and three fetuses from four distinct families. Affected individuals presented with short stature and mesomelic limb shortening, in those for whom X-rays were available. Additional common anomalies included a bell-shaped chest, short neck, and prominent deltoid tuberosities. Radiological examinations revealed characteristic findings, including metaphyseal irregularities, pseudoepiphyses of the fingers, flared iliac wings, and a wavy appearance of the ribs. Some individuals also exhibited early degenerative joint changes, mild scoliosis, and vertebral wedging, particularly affecting the thoracolumbar spine. Dental anomalies were a prominent feature, with brownish discoloration of the teeth, enamel hypoplasia, and signs of dentinogenesis imperfecta in patients for whom dental and orthopanoramic examination were performed. The three fetuses had prenatal findings detected via ultrasound suggestive of skeletal dysplasia, including severe shortening of the long bones, curved femurs, and thoracic hypoplasia. All affected individuals had normal developmental milestones and no major involvement of other organ systems. A summary of all clinical and radiological findings including review of previously reported cases is presented in Extended Data Figure 4 and Supplementary Table 5.

### SILAC secretome analysis

For the quantitative analysis of the TANGO1 or HSP47-dependent secretome, SILAC (stable isotope labeling by amino acids in cell culture) experiments were performed with four independent biological replicates, including two independent TANGO1 KO or HSP47 KO clones as described before ^16^. Note that in secretome studies, full FBS-containing culture media were replaced with media lacking serum for 24h before sample collection, thus removing the highly abundant serum proteins from the medium before protein identification to increase the probability to detect low abundant secreted proteins.

In brief, wild-type and TANGO1 or HSP47 knock-out cells were pulse-labeled at 70% confluency. Cells were pre-incubated in SILAC medium (Met/Arg/Lys-free DMEM/F12; #AES-0423, Athenaes), supplemented with 1 mM sodium pyruvate (11360070, Thermo Fisher Scientific), 200 mg/L L-proline (#P0380, Sigma-Aldrich), and 10% dialyzed FBS (dFBS; #AES-0427, Athenaes) for 30 min, to deplete intracellular Methionine, Arginine, and Lysine. Cells were then labeled for 24 h in SILAC medium supplemented with 150 μg/ml ascorbic acid-2-phosphate, 1 mM sodium pyruvate (11360070, Thermo Fisher Scientific), 200 mg/L L-proline (#P0380, Sigma-Aldrich), 100 μM azide-group-containing methionine analog azidohomoalanine (AHA; 63669AS, Anaspec), and either heavy (146 μg/ml Lys-8 (^13^C6,^15^N2) / 84 μg/ml Arg-10 (^13^C6, ^15^N4)) or medium isotopes (146 μg/ml Lys-4 (^2^H) / 84 μg/ml Arg-6 (^13^C6)). All isotope-labeled amino acids were purchased from Silantes (#211103913, #201203902, #211603902, #201603902). Supernatants of wild-type heavy isotope-labeled and knock-out medium isotope-labeled cultures were combined, dead cells were removed by centrifugation (5 min, 1000 x g, 4°C), and proteins were concentrated to approximately 250 μl, using Amicon ultra-15, PLBC Ultracel-PL 3 kDa cut-off concentrator tubes (#Z740199, Sigma-Aldrich). Cells were lysed in 850 μL of urea lysis buffer provided with the Click Chemistry Capture kit (CLK-1065, Jena Bioscience), according to the manufacturer’s protocol. Supernatants and cell lysates were individually covalently coupled to 200 μL pre-washed (2x with 1 mL HPLC H_2_O each) alkyne agarose (CLK-1032-2, Jena Bioscience), rotating overnight at RT. Alkyne agarose-bound samples were spun down (5 min, 2000 x g, 4°C), washed once with 1 mL HPLC water, and then reduced with 5 mM DTT (15 min, 70°C), alkylated with 40 mM CAA (30 min, RT), transferred to columns (supplied with the kit) and extensively washed sequentially with SDS wash buffer (supplied with the kit), 100 mM Tris-HCl pH 8.0 / 8 M urea buffer, 20% isopropanol, and 20% acetonitrile (20 mL each). Proteins were then digested overnight with 1 μg trypsin (#37286, Serva) and 0.5 μg LysC (#129-02541, FUJIFILM Wako Chemicals) in a heated shaker (37°C, 800 rpm). Digestion was stopped by adding 0.5% formic acid to the reaction, and the peptides were subjected to StageTip purification, before injection into the mass spectrometer.

### Gene Ontology analysis

Gene Ontology (GO) and pathway enrichment analysis were performed using the Database for Annotation, Visualization and Integrated Discovery (DAVID) tool ^48^, as described previously ^16^. In brief, for the analysis of the TANGO1 and HSP47-dependent secretome, proteins whose intensity decreased robustly and significantly (log_2_FC > 1, p < 0.05) in the supernatants of TANGO1 KO versus control WI-26 cells, (log_2_FC > 0.6, p < 0.05) HSP47 KO versus control WI-26 were used for GOTERM_CC_FAT, GOTERM_BP_FAT, KEGG_PATHWAY, GOTERM_MF_FAT, and GOTERM_PFAM analyses (Table S1 and S2). The human proteome was used as reference list. The data were visualized in a cell plot, generated using DAVID and the associated Flaski apps (https://flaski.age.mpg.de, developed and provided by the MPI-AGE Bioinformatics core facility ^49^ using 8 selected non-redundant significant CC_GO terms for the secretome.

The full list of proteins detected in the secretome experiment was used for generating the Volcano plots. Proteins whose secretion changes significantly between TANGO1 KO or HSP47 KO and control cells are shown as grey dots, non-significant hits as white dots. Proteins corresponding to the GO term ‘GO:0062023 collagen-containing extracellular matrix’ are shown as red dots.

### Recombinant protein production in HEK293FT cells

TST (Twin-Strep-Tagged) fusion proteins were produced from pITR-TTP-TANGO1 SH3L/MOTH vectors containing the cDNA coding for the SH3L/MOTH domain of TANGO1 WT, C38Y, F63C or Y75R, as well as murine col6a1, col6a2, col6a3 and col6a3 fragments in stable HEK293FT using the sleeping beauty transposon system as described earlier. The expression of TST-SH3L/MOTH WT, C38Y, F63C and Y75R was induced with 1 μg/ml Doxycycline for 96 h in serum-free DMEM/F12 + L-Glutamine. TST fusion proteins were purified using StreptactinXT 4Flow high-capacity resin (#IBA-2-5030-002, IBA Lifesciences) used for subsequent affinity isolation assays or eluted using 100 mM Tris/HCl pH 8.0, 150 mM NaCl, 1 mM EDTA, 50 mM Biotin for SPR measurements. The recombinant expression and affinity purification of recombinant proteins spanning fibrillin-1 and fibrillin-2 was previously described ^36,37^.

### TST affinity precipitation assay

For TST affinity precipitation assays, three 10 cm dishes of WI-26 cells, cultured with 150 μg/ml ascorbic acid-2-phosphate for 24 h were lysed in 1 ml Triton lysis buffer (as described above) each, and 500 μl of the lysates were incubated with equal amounts of recombinant TST-fusion proteins bound to Streptactin XT resin overnight at 4 °C with overhead rotation. The next day, samples were washed 4 x with Triton lysis buffer, boiled in 1x Laemmli sample buffer (5 min, 95°C), and separated by SDS-PAGE. Input samples (50 μl) were collected before pre-clearing, SDS sample buffer with or without DTT (1x final concentration) was added to the lysates, samples were boiled and analyzed by immunoblotting.

### SPR measurements

SPR experiments were performed as described previously ^50^ using a BIAcore 2000 system (Cytiva, Marlborough, MA). CM5 chips (#BR100012, Cytiva, Marlborough, MA) were calibrated with running buffer HBS-EP (0.01 M HEPES, pH 7.4, 0.15 M NaCl, 3 mM EDTA, 0.005% (v/v) surfactant P20) (#BR100188, Cytiva, Marlborough, MA). To narrow down the TANGO1 SH3L/MOTH-binding site within the different ECM proteins and ECM protein fragments, a CM5 chip was calibrated with HBS-EP buffer and TANGO1 SH3L/MOTH WT or Y75R mutant was immobilized via NHS/EDC amine coupling (#BR100050, Cytiva, Marlborough, MA) at 1000 RUs. Full length ECM proteins or fragments were injected in a dilution series from 0 to 320 nM in HBS-EP buffer.

### Molecular modeling

The models of TANGO SH3 in complex with COL6A3 were calculated using AlphaFold 3 ^51^ and conservation was analyzed with the ConSurf Web Server ^52,53^. Figures were prepared using ChimeraX ^54^.

### Statistical analysis

Statistical analysis and presentation of quantification data was performed using GraphPad Prism (version 10). Data in all graphs are shown as mean ± SD. Significance for the indicated pairwise comparisons was calculated using one-way Anova. Sample sizes (n) and significance values are indicated in figure legends (* p < 0.05, ** p < 0.01, *** p < 0.001, **** p < 0.0001).

All findings were reproducible over multiple independent experiments, within a reasonable degree of variability between replicates. For most experiments, at least three independent replicates were performed. The sample size for microscopy analyses (number of individual cells used for quantification) is provided in the respective figure legends. No statistical method was used to predetermine sample size, which was determined in accordance with standard practices in the field. No data were excluded from the analyses. The experiments were not randomized, and the investigators were not blinded to allocation during experiments and outcome assessment.

## Supporting information

Secretome Dataset TANGO1 KO

Secretome Dataset HSP47 KO

Antibodies used in this study

Oligonucleotides used in this study

Clinical Table MIA3/TANGO1 variants

## Acknowledgements

We dedicate this manuscript to Dr. Markus Plomann, who sadly passed away in April 2024, during preparation of this manuscript. We thank Gloria Kyrilla and Carolin Poremba for technical support and all members of the Demetriades lab for critical discussions; the MPI-AGE FACS & Imaging Core Facility for support with confocal microscopy; and the MPI-AGE Proteomics Core Facility for support with the proteomics experiments described in this study. We thank the Institute for Biochemistry II, the Lemberg lab for helpful discussions and Marius Lemberg for critical reading of the manuscript. CD is funded by the European Research Council under the European Union’s Horizon 2020 research and innovation programme (grant agreement No 757729), and by the Max Planck Society. JN acknowledges support by the Center for Molecular Medicine Cologne (CMMC) for funding through the Individual Project Funding Program (Project C10) and the Career Advancement Program (Project CAP31). Parts of this work were supported by the Deutsche Forschungsgemeinschaft (DFG, German Research Foundation) through the Research Unit Grant FOR2722 (NU 467/1-1, SCHI 1627/2-2, SE 1115/4-2, and DE 3170/1-1; Project No 384170921) to JN, AS, GS, and CD, respectively; the DFG Grant NU 467/2-1 (project No 564334266) to JN; the DFG grant KR 558/17-1 (project No 468236352) to TK; and the DFG Grants TRR259/B09 (project No 397484323), and SE1115/7-1 (project No 468236352) to GS. VC is member of the European Reference Network on Rare Congenital Malformations and Rare Intellectual Disability ERN-ITHACA, funded by the European Union (grant agreement No. 101156387). KEH and MP-P are members of the European Reference Network on BOne Disorders (ERN BOND) funded by the European Union (grant agreement No. 101157116). ABZ is member of the ERN CRANIO by the European Union (grant agreement No. 101156275). Models in figures created with BioRender.com.

## Author Contributions

Experimental work: JN, SL, ST, FO, AS, assisted by SW and MT; data analysis: JN, SL, CD; Recombinant protein production: MT, AS, SL; structural analysis: ST, DK; patient cohorts: AA, VC, RM, MZ, MVP, KEH, ABZ, AO, TANGO1 consortium; project design, conceptualization: JN, AA, CD; supervision: JN, MP, CD; funding acquisition: JN, TK, MK, AS, GS, CD; figure preparation: JN, SL, AO, ST, ABZ; manuscript draft: JN, with contributions from AA, BE, TK, DK, MK, AS, ST, MVP, ABZ, KEH, GS, and CD. All authors and members of the TANGO1 consortium approved the final version of the manuscript and agree on the content and conclusions.

## Data availability

All data needed to evaluate the conclusions in the paper are present in the paper and/or the Supplementary Material. The data that support the findings of this study (uncropped immunoblots, microscopy pictures) are available from the corresponding authors upon reasonable request.

## Code availability

No code was generated in this study.

## Additional Information

Supplementary Information (Extended Data Figures 1-7 and Supplementary Tables 1-5) is available for this paper.

