## Supplementary Information for "A conserved cargo recognition mechanism establishes TANGO1 as a selective extracellu lar matrix cargo receptor"

Supplementary Tables 1-5, Extended Data Figures 1-7, and Supplementary Notes

### Supplementary Tables

Supplementary Table 1. TANGO1-dependent secretome.

Supplementary Table 2. HSP47-dependent secretome.

Supplementary Table 3. List of primary antibodies used in this study.

Supplementary Table 4. List of DNA oligonucleotides used in this study.

Supplementary Table 5: Clinical information for the affected individuals carrying *MIA3* variants

### Extended Data Figures

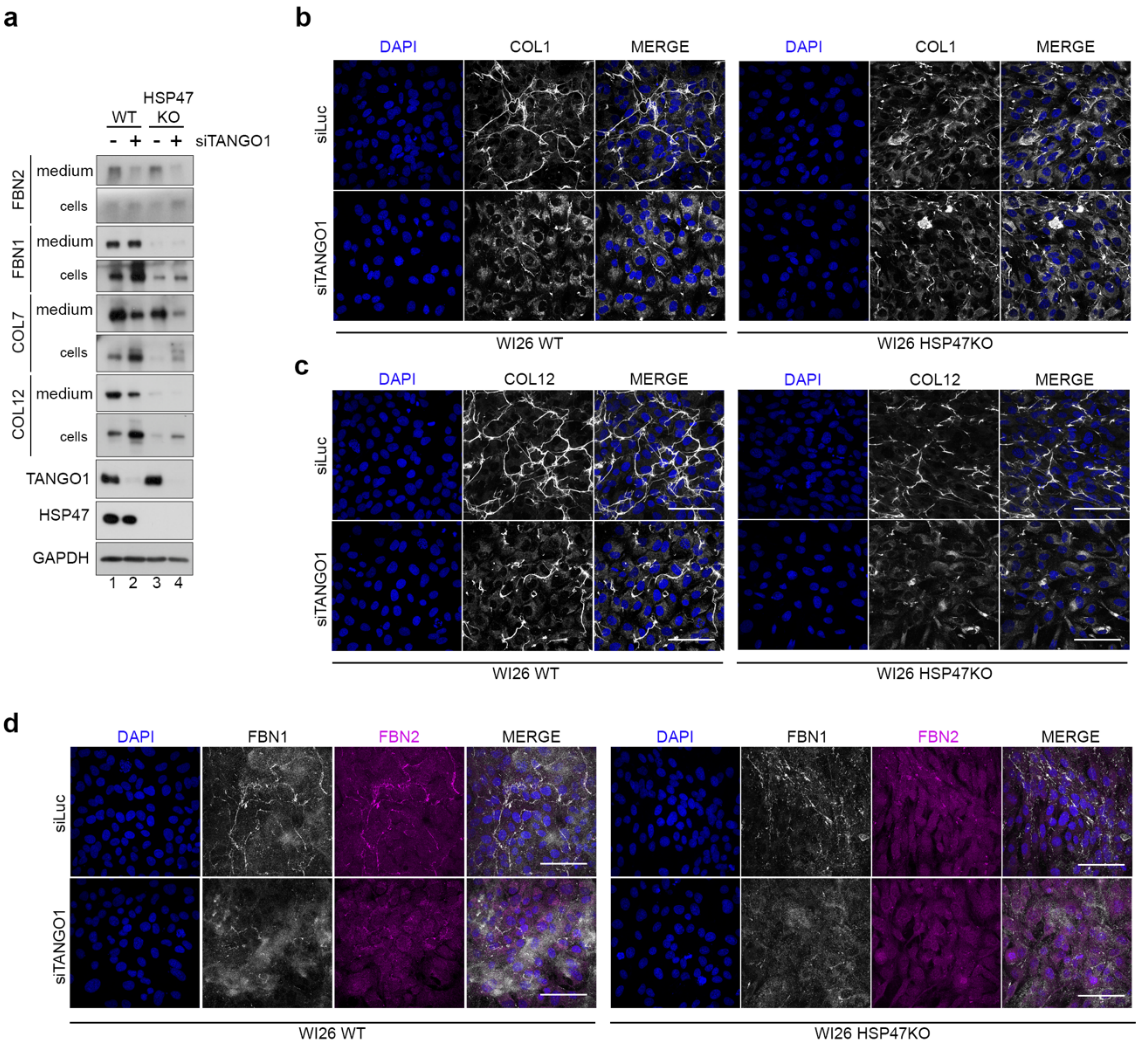


**Extended Data Figure 1. Secretion of selective bulky cargo is still TANGO1-dependent in the absence of HSP47. Related to Figure 1.**

**(a)** Western blot analysis of cell lysates (cells) and supernatants (medium) derived from WT and HSP47 KO WI26-SV40 cells transfected with siLuc (-) or siTANGO1 (+) using antibodies as indicated.

**(b-d)** Immunofluorescence analysis of WT and HSP47 KO WI26-SV40 cells transfected with siLuc or siTANGO1 using type I collagen (COL1) (b), type XII collagen (COL12) (c), and fibrillin 1 & 2 (FBN1, FBN2) (d) antibodies. Nuclei are stained with DAPI. Scale bars, 10 µm. n = 3 independent experiments.

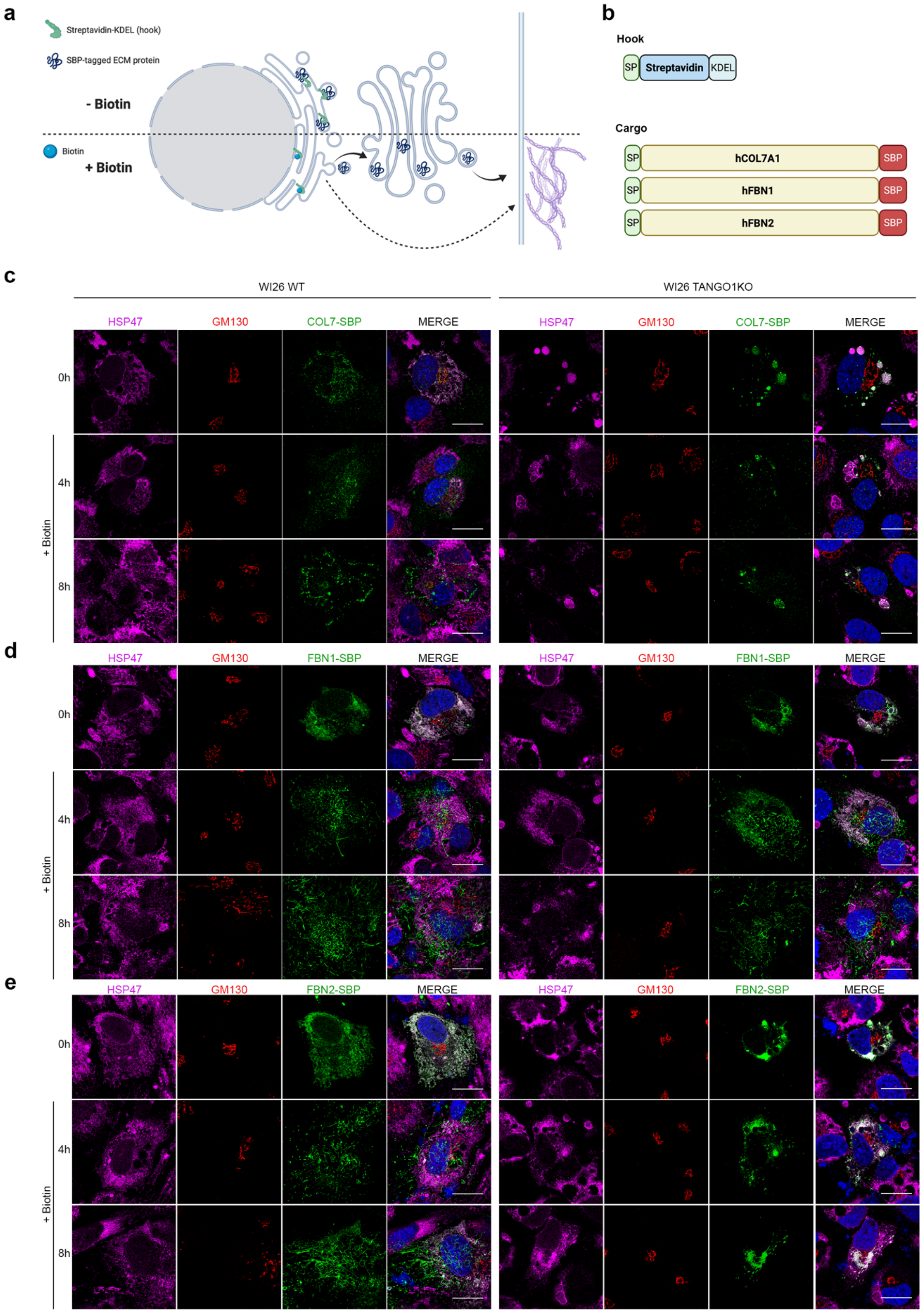


**Extended Data Figure 2. SBP-tagged COL7A1 and FBN2, but not FBN1 are retained in ER of TANGO1 KO cells in a RUSH assay. Related to Figure 1.**

**(a)** Scheme of experimental setup for RUSH analyzing ECM proteins.

**(b)** Scheme of constructs used in this assay. SP= Signal Peptide, SBP= Streptavidin-binding peptide.

**(c)** Immunofluorescence analysis of WT or TANGO1 KO WI-26 cells transiently expressing streptavidin-binding peptide (SBP)-tagged type VII collagen (COL7A1) at the indicated time points following biotin addition.

**(d)** Immunofluorescence analysis of WT or TANGO1 KO WI-26 cells transiently expressing streptavidin-binding peptide (SBP)-tagged Fibrillin-1 (FBN1) at the indicated time points following biotin addition.

**(e)** Immunofluorescence analysis of WT or TANGO1 KO WI-26 cells transiently expressing streptavidin-binding peptide (SBP)-tagged Fibrillin-1 (FBN1) at the indicated time points following biotin addition.

For all stainings, GM130 was used as a Golgi marker, HSP47 as an ER marker. Nuclei stained with DAPI. Scale bars, 10 µm. n = 3 independent experiments.

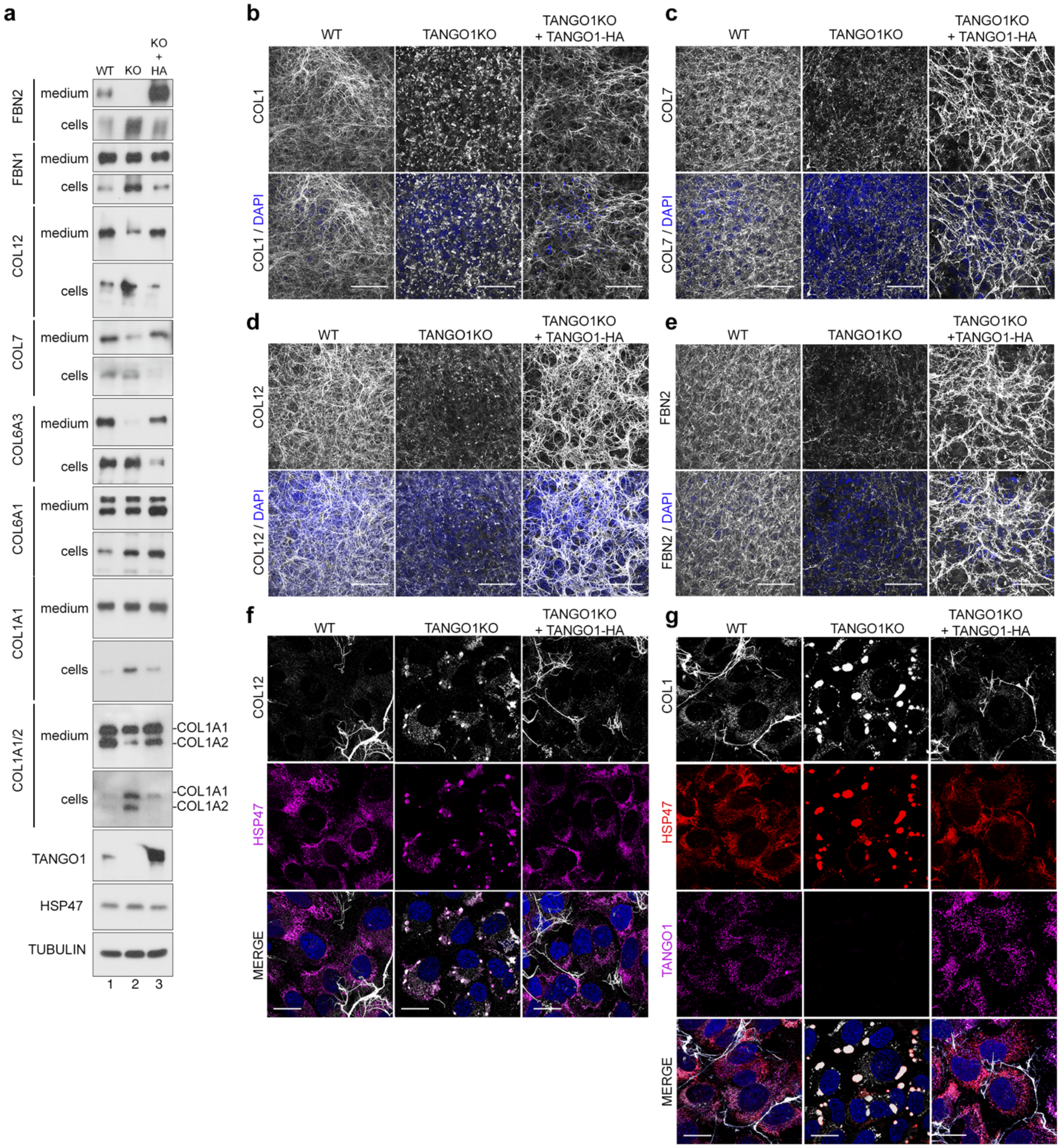


**Extended Data Figure 3. Stable expression of TANGO1-HA fully reverses the ER retention phenotype of TANGO1 KO cells. Related to Figures 1-2.**

**(a)** Western blot analysis of cell lysates (cells) and supernatants (media) of WT, TANGO1 KO (KO) and TANGO1 KO cells, cultured for 24 h in the presence of ascorbic acid, stably reconstituted with TANGO1-HA (KO+HA) with the indicated antibodies.

**(b-e)** Immunofluorescence analysis of WT, TANGO1 KO and TANGO1 KO cells reconstituted with TANGO1-HA, cultured in the presence of ascorbic acid for 7 days, stained for type I collagen (COL1, b), type VII collagen (COL7, c), type XII collagen (COL12, d) and fibrillin-2 (FBN2, e). Nuclei stained with DAPI. Scale bars, 50 µm. n = 3 independent experiments.

**(f-g)** Immunofluorescence analysis of WT, TANGO1 KO and TANGO1 KO cells reconstituted with TANGO1-HA, cultured in the presence of ascorbic acid for 2 days, stained for COL12 and HSP47 (f) and COL1, HSP47 and TANGO1 (g). Nuclei stained with DAPI. Scale bars, 10 µm. n = 3 independent experiments.

a

**contact the corresponding author to request access to the pedigrees**

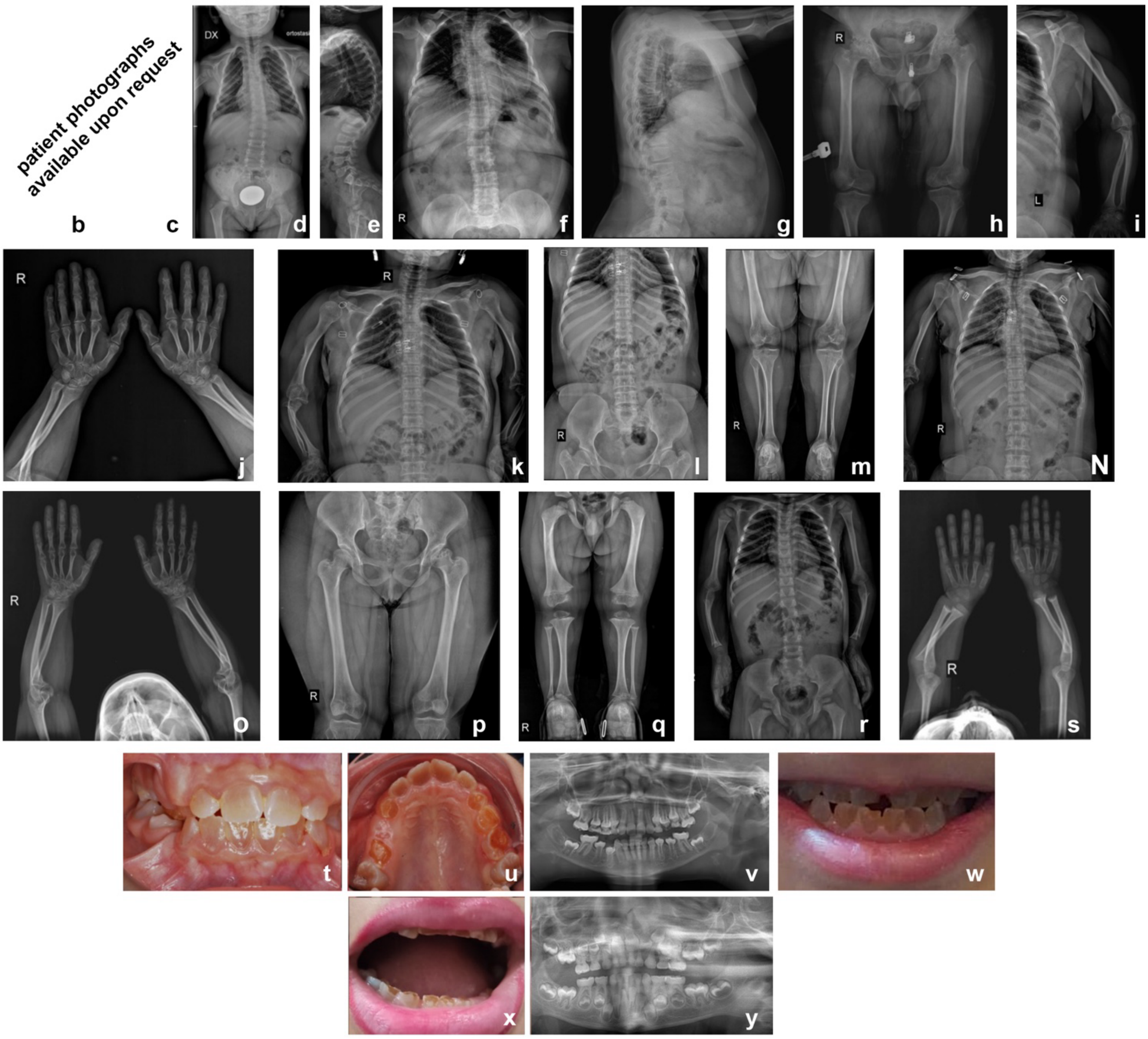

**Extended Data Figure 4. Genetic and phenotypic characteristics of individuals with biallelic *MIA3* variants. Related to Figure 4.**

**(a-y)** Pedigree of families 1-4 (a) showing the affected individuals (shaded). Genotypes of tested individuals are indicated under the symbols. + represents the reference allele. Lateral and front view images of individual II:2 from Family 1 at age range 1-5 and 16-20 years, respectively, show lumbar lordosis (b) and short stature (c). a-c contain sensitive information and are only available upon reasonable request. Skeletal surveys show common and recurrent radiographic features among affected individuals, including mesomelia, prominent deltoid tuberosities, relatively broad medial ends of the clavicles and downward sloping mildly wavy ribs in individuals V:3 (k, l, m), IV:3 (f, g, h, i, j) , V:4 (n, o, p) and V:2 (q, r, s) of Family 2. Additional features seen in older patients of Family 2 (IV:3, V:3, V:4) include short femoral necks and flattening, degenerative disease with narrow joint spaces and erosions/cysts of the femoral heads (h, l, p). Less common and non-specific features include mild thoracolumbar scoliosis and exaggerated lumbar lordosis/dorsal kyphosis in subject II:2 of Family 1 (d, e) and mild cupping of the metaphyses of the terminal phalanges and pseudoepiphyses of the thumb in subject V:2 of Family 2 (s). Clinical intraoral pictures (t, u) and panoramic radiograph (v) of subject II:2 of Family 1 at age range 5-10 years old demonstrate typical features of dentinogenesis imperfecta, apparent in both the primary and permanent dentitions with grey amber brownish translucent teeth, and enamel wear clearly visible on primary maxillary teeth. Panoramic radiograph (v) shows bulbous crowns, cervical constriction, short roots, thin enamel, evolution from thin root dentin walls and large pulp chambers (as seen on erupting premolars) towards progressive disappearance and obliteration of pulp spaces (lower incisors). Clinical intraoral pictures of subject V:2 (w) and V:4 (x) and panoramic radiograph of subject V:2 (y) of Family 2 at the age range of 0-5 years old also shows typical dentinogenesis imperfecta.

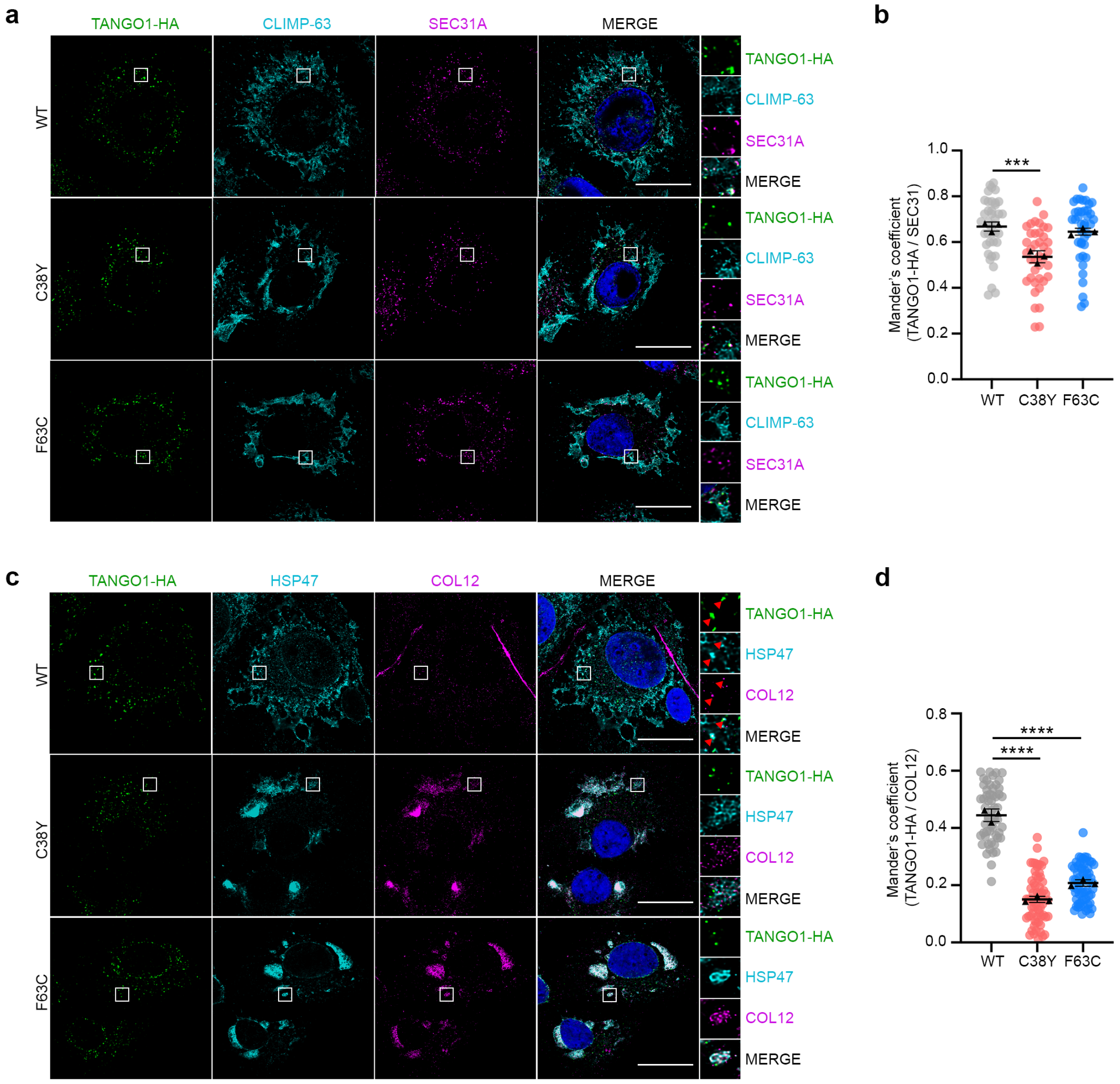


**Extended Data Figure 5. Stable reconstitution of TANGO1-HA patient variants resembles correct localization to ER-exit sites but show reduced colocalization with cargo proteins. Related to Figure 5.**

**(a)** Immunofluorescence analysis of TANGO1 KO cells reconstituted with TANGO1-HA WT, C38Y and F63C, cultured in the presence of ascorbic acid for 2 days, stained for SEC31, CLIMP63 and HA. Magnified insets shown to the right.

**(b)** Mander's correlation coefficient of TANGO1-HA and SEC31 quantified from (A). $p<0.001$, one-way ANOVA. Data in graphs shown as mean ± SD.

**(c)** Immunofluorescence analysis of TANGO1 KO cells reconstituted with TANGO1-HA WT, C38Y and F63C, cultured in the presence of ascorbic acid for 2 days, stained for type XII collagen (COL12), HSP47 and HA. Magnified insets shown to the right, red arrowheads highlighting colocalization of three channels.

**(d)** Mander's correlation coefficient of TANGO1-HA and COL12 quantified from (C). **** $p<0.0001$, one-way ANOVA. Data in graphs shown as mean ± SD. Scale bars, 10 μm. n = 3 independent experiments.

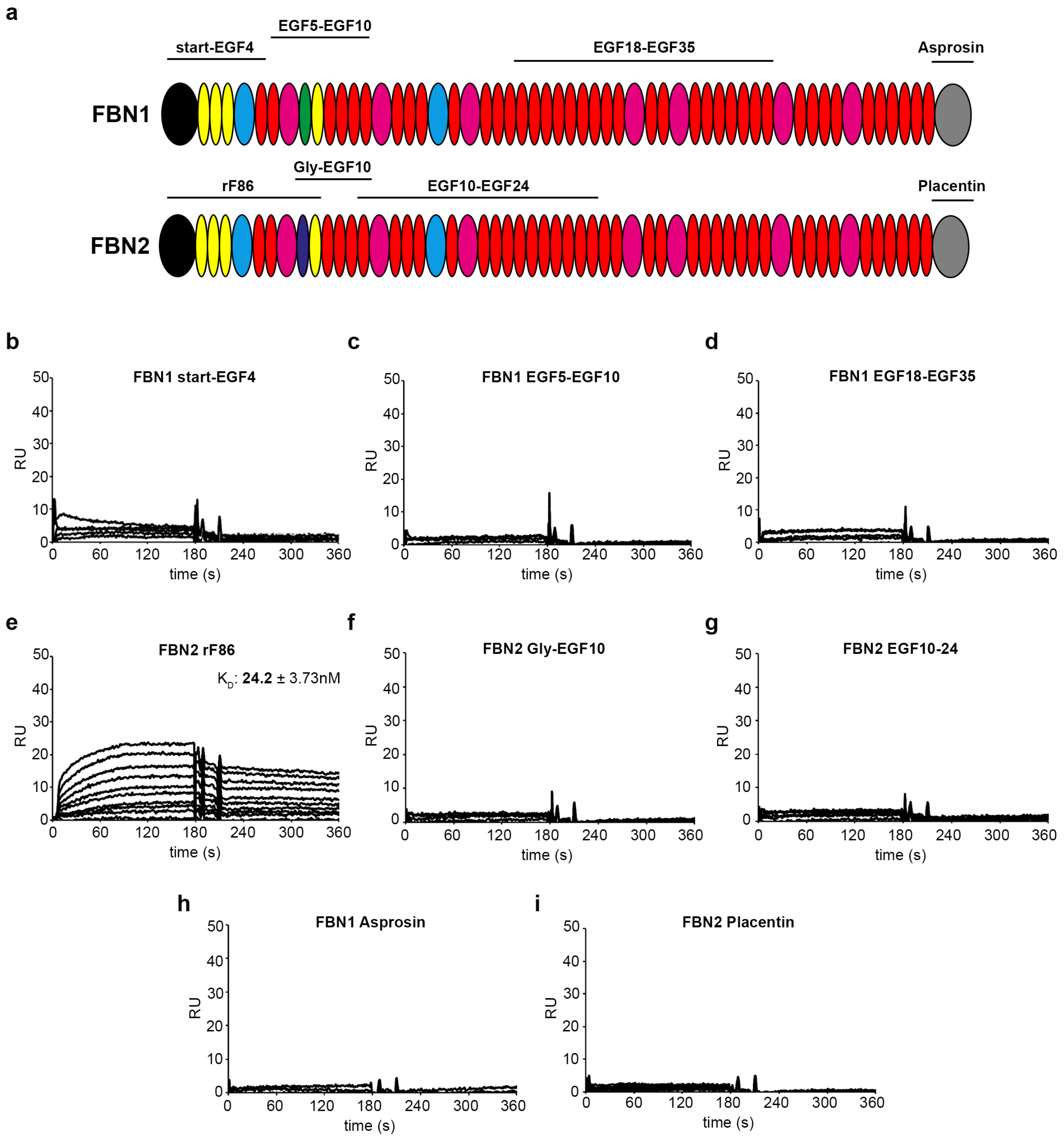


**Extended Data Figure 6. TANGO1 selectively binds to the N-terminus of FBN2.**

**(a)** Domain structures of human fibrillin-1 (FBN1) and fibrillin-2 (FBN2). Protein fragments used for SPR measurements are marked respectively.

**(b-i)** SPR sensorgrams obtained either of the interaction between TANGO1-MOTH domain (immobilized on the sensor chips in similar quantities) and human FBN1 (b,c,d,h) or FBN2 fragments (e,f,g,i) (flown over as soluble ligands) in a dilution series from 0 to 320 nM. Ligand binding to the immobilized proteins on the chip is shown as response units on the y-axis. n = 3 independent experiments.

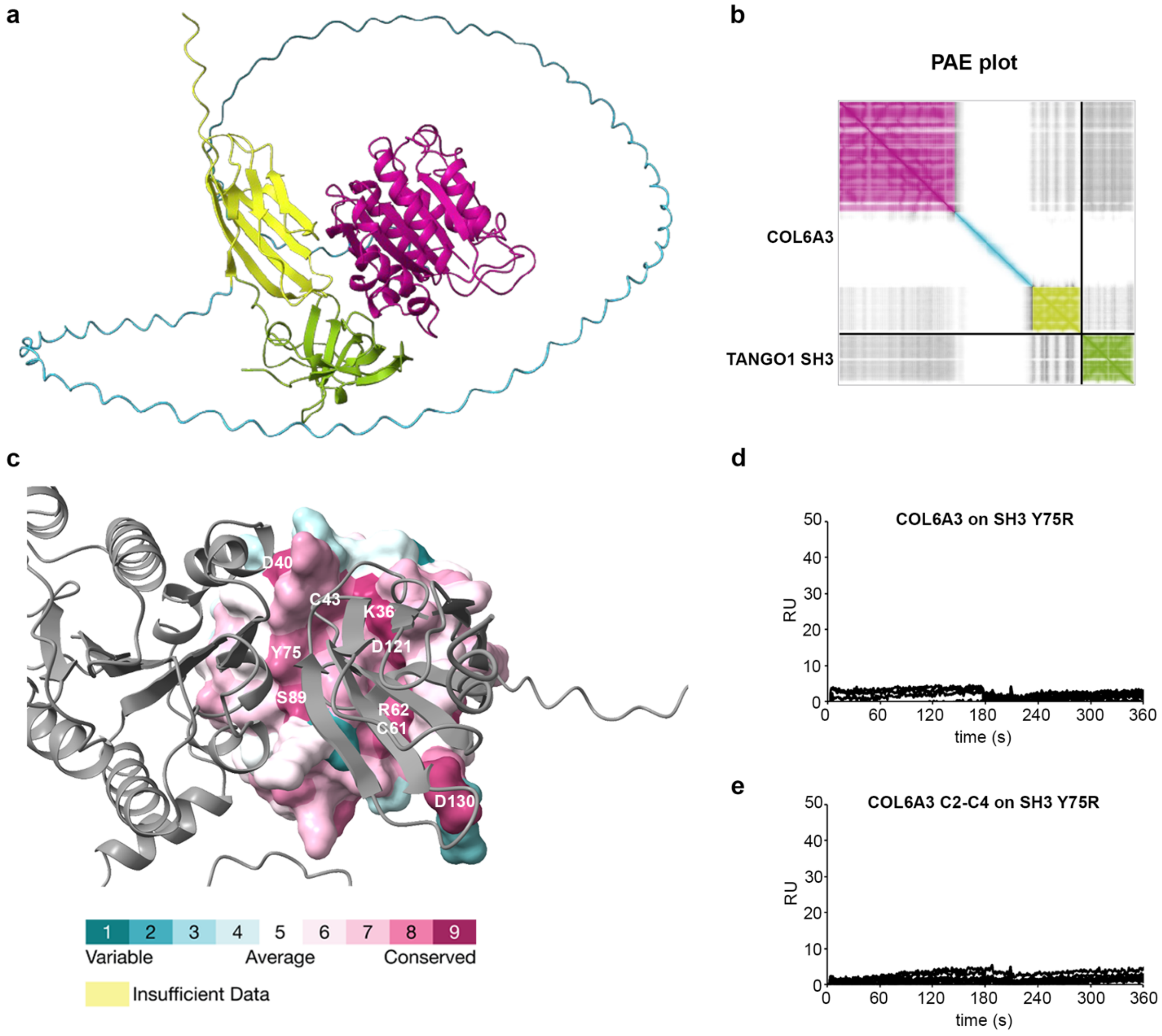


**Extended Data Figure 7. Highly conserved Tyr75 in the MOTH domain of TANGO1 is involved in selectively recognizing COL6A3 as a cargo protein. Related to Figures 6-7.**

**(a-b)** Accuracy of the AlphaFold3 models. (a) TANGO MOTH in complex with COL6A3 colored by AlphaFold3 recognized domains. (b) The interdomain model accuracy is represented by the Predicted Alignment Error (PAE) plot, colored by domains as in (A).

**(c)** Interface of COL6A3 with TANGO1 MOTH, COL6A3 shown in surface representation colored by coulomb potential and TANGO MOTH shown as cartoon in grey.

**(d-e)** SPR sensorgrams obtained either of the interaction between TANGO1 MOTH domain Y75R mutant (immobilized on the sensor chips in similar quantities) and human COL6A3 **(**d) or COL6A3 C2-C4 fragment (e) (flown over as soluble ligands) in a dilution series from 0 to 320 nM. Ligand binding to the immobilized proteins on the chip is shown as response units on the y-axis. n = 3 independent experiments.

### Supplementary Notes

### TANGO1 Consortium Information (in alphabetical order)

Andrea Accogli[4,5,6], Anna Elsa Maria Allegri[26], Valeria Capra[27], Claudia Capurro[26], Patrizia De Marco[28], Francisca Diaz Gonzalez[29], Natascia Di Iorgi[26,30], Mohammad Hamid[31], Mohamad Ghasem Hanafi[32], Michele Iacomino[27], Nicola Laffi[26], Reza Maroofian[12], Aleš Maver[33], Manuel Parrón-Pajares[34], Siavash Sadeghian[35], Vincenzo Salpietro[36,37], Alireza Sedagahat[38,39], Marcello Scala[27,30], Gholamreza Shariatia[38,40], Claudia Toledo-Pacheco[41], Mateja Vinkšel Prah[14], Mina Zamani[12,13], Federico Zara[27,30], Jawaher Zeighami[38]

[26] Department of Pediatrics, IRCCS Istituto Giannina Gaslini, Genoa 16147, Italy.

[27] Genomic and Clinical Genetics Unit, IRCSS Istituto G. Gaslini, Genoa, Italy.

[28] Medical Genetics Unit, IRCCS Istituto Giannina Gaslini, Genoa, Italy.

[29] Institute of Medical and Molecular Genetics (INGEMM), Hospital Universitario La Paz, IdiPAZ, UAM, Madrid, Spain.

[30] Department of Neuroscience, Rehabilitation, Ophtalmology, Genetics, Maternal and Child Health, University of Genoa, Genoa 16142, Italy.

[31] Department of Molecular Medicine, Biotechnology Research Center, Pasteur Institute of Iran, Tehran, Iran

[32] Department of Radiology, School of Medicine, Ahvaz Jundishapur University of Medical Sciences, Ahvaz, Iran

[33] University Medical Centre Ljubljana, Clinical Institute of Genomic Medicine, Ljubljana, Slovenia

[34] Department of Radiology, Hospital Universitario La Paz, Madrid, Spain.

[35] Department of Orthopedic Surgery, Arak University of Medical Sciences, Arak, Iran

[36] Pediatrics Unit-Department of Biotechnological and Applied Clinical Sciences, University of L'Aquila, 67100 L'Aquila, Italy

[37] European Brain Research Institute "Rita Levi-Montalcini" Viale Regina Elena, Rome, Italy.

[38] Narges Medical Genetics and Prenatal Diagnosis Laboratory, Kianpars, Ahvaz, Iran

[39] Diabetes Research Center, Health Research Institute, Ahvaz Jundishapur University of Medical Sciences, Ahvaz, Iran

[40] Department of Medical Genetics, Faculty of Medicine, Ahvaz Jundishapur University of Medical Sciences, Ahvaz, Iran

[41] Department of Genetics, Hospital Universitario de Toledo, Toledo, Spain.
